# MIND-NL diet adherence moderates the relation of low-grade systemic inflammation with neuroinflammation and cognitive functioning: an exploratory cross-sectional study in older adults

**DOI:** 10.64898/2026.01.21.26344505

**Authors:** Lianne B. Remie, Mark R. van Loenen, Mara P.H. van Trijp, Ilke G.S. de Lange, Yannick Vermeiren, Jurriaan J. Mes, Nicolaas A. Puts, Joukje M. Oosterman, Esther Aarts

**Affiliations:** Radboud University, Donders Institute for Brain, Cognition and Behaviour, Nijmegen, the Netherlands; Division of Human Nutrition and Health, Wageningen University & Research, Wageningen, the Netherlands; Wageningen Food and Biobased Research, Wageningen University & Research, Wageningen, the Netherlands; Department of Forensic and Neurodevelopmental Sciences, Institute of Psychiatry, Psychology, and Neuroscience, King’s College London, London, UK; MRC Centre for Neurodevelopmental Disorders, King’s College London, London, UK

**Keywords:** MIND diet, dietary pattern, systemic inflammation, neuroinflammation, cognition, intestinal-barrier permeability, ageing, cross-sectional

## Abstract

**Background:** Observational studies have linked high adherence to the “Mediterranean-Dietary Approaches to Stop Hypertension Intervention for Neurodegenerative Delay” (MIND) diet to improved cognitive functions in older adults. The underlying peripheral and central mechanisms of this association remain poorly understood, although multiple nutrients in the MIND diet are known for their anti-inflammatory effects. Therefore, we explored the cross-sectional relation between MIND diet adherence (Dutch version), systemic inflammation, neuroinflammation, and cognitive functioning in older adults. In addition, we examined the role of intestinal barrier permeability in MIND diet associations with (neuro)inflammation.

**Methods:** We included 88 older adults (60-75 year) at risk of cognitive decline. MIND-NL diet adherence was assessed using a food frequency questionnaire. Systemic inflammation (C-reactive protein levels, white blood cell-counts and neutrophil-to-lymphocyte ratio) and intestinal barrier permeability (lipopolysaccharide-binding protein, zonulin, and lipopolysaccharide) markers were measured in blood. Neuroinflammation-associated metabolites (myo-inositol, choline and creatine) were measured in the dorsolateral prefrontal cortex with proton magnetic resonance spectroscopy (^1^H-MRS). Cognitive functioning was assessed with a neuropsychological test battery.

**Results:** Linear models showed that both MIND diet adherence and systemic inflammation did not predict neuroinflammation or cognition independently. However, MIND diet adherence significantly moderated the relation between systemic inflammation and neuroinflammation (β=-0.11, *p*=0.04) as well as between systemic inflammation and cognition (β=0.044, *p*=0.02). Specifically, in individuals with lower MIND diet adherence (identified as scores ≤7), systemic inflammation was positively related to neuroinflammation, and negatively to cognition. Similarly, MIND diet adherence significantly moderated the relation between intestinal barrier permeability and neuroinflammation (β=-0.17, *p*=0.05). Finally, within participants with lower MIND adherence (median split at ≤8.75), systemic inflammation mediated the relation between the intestinal barrier permeability and neuroinflammation (β=0.427 [0.072; 0.891], *p*=0.04).

**Conclusion:** Our findings suggest that higher MIND diet adherence might protect against the detrimental effect of systemic inflammation on neuroinflammation and cognitive functioning. Moreover, we demonstrated that greater adherence to the MIND diet may specifically protect against the systemic inflammation-mediated relationship between intestinal barrier permeability and neuroinflammation. These findings should be confirmed in randomised controlled trials.

## Background

In our ageing world population, focus on prevention and delay of cognitive decline is of utmost importance. The World Health Organization recognizes lifestyle modification as holding the greatest potential among preventive approaches (1). Diet, a major lifestyle domain, is highly modifiable and has been identified as an important factor associated with cognitive decline (2,3). However, the peripheral and central mechanisms by which diet may affect the underlying processes of cognitive decline in humans remain unclear.

The Mediterranean-Dietary Approaches to Stop Hypertension (DASH) Intervention for Neurodegenerative Delay (MIND) diet integrates features from both the Mediterranean and DASH diets (4), such as berries and green leafy vegetables. Importantly, large observational cohort studies have linked MIND diet adherence to decreased incidence rates of Alzheimer’s Disease (AD) and better cognitive performance in healthy older adults in cross-sectional analyses, specifically within the global cognition and episodic memory domains (5). However, longitudinal evidence for protective effects against cognitive decline remains limited (5). Moreover, randomized controlled trials (RCTs) that investigated a MIND diet intervention compared to a control diet show inconsistent effects on cognition, with one 3-month trial in middle-aged obese women demonstrating significant improvements in cognitive functioning (6) while another 3-year trial in older adults found no such effects (7). To advance research in this area, it is important to elucidate the underlying mechanisms through which MIND diet components may influence cognitive brain ageing.

The MIND diet is rich in vitamins, fibers, omega-3 fatty acids, and antioxidants such as polyphenols (8). All of these nutrients could exert anti-inflammatory effects and reduce oxidative stress, either directly (e.g. omega-3 fatty acids) or indirectly via the gut microbiome (e.g. fibers being fermented into short-chain fatty acids) (9). Moreover, food components with established pro-inflammatory effects (e.g., saturated fats, processed meats) are discouraged within the MIND diet (8,9). Collectively, these insights point to inflammation modulation as a potential key mechanistic pathway underlying MIND diet’s neuroprotective capacity against cognitive decline, especially given the pivotal roles of both peripheral- and central inflammatory processes in cognitive ageing (10).

Ageing, combined with prolonged exposure to lifestyle-related stressors, can shift the peripheral immune system towards a more pro-inflammatory state (11). As a result, chronic low-grade systemic inflammation is commonly seen in older adults, and characterized by a subtle and sustained elevation in circulating immune markers, such as C-reactive protein (CRP), interleukin (IL)-6, and tumor necrosis factor (TNF)-α (12). The quantification of low-grade systemic inflammation in previous studies has often relied on individual biomarkers, with CRP levels (3-10 mg/L) serving as the gold standard (13). However, composite inflammatory indices combining CRP with white blood cell (WBC) parameters, such as the INFLA score, offer a more comprehensive evaluation of inflammatory processes and demonstrate increased sensitivity in detecting low-grade inflammation (14,15). Age-related low-grade systemic inflammation is suggested to arise via multiple underlying pathways. Among these, the intestinal epithelial barrier is considered a key shaper of the immune system, as it houses a substantial proportion of the body’s immune cells (16). Importantly, increased intestinal barrier (IB) permeability as a result of lifestyle-related stressors (such as diet) has been associated with increased systemic inflammation (17).

Limited research has examined the association between MIND diet adherence and systemic inflammation markers in older adults. Two recent observational studies in older adults showed that higher MIND diet adherence was associated with lower circulating inflammatory markers (including CRP and WBC counts) and better cognitive performance (18,19), with systemic inflammation mediating the diet-cognition relationship (18). One of these, a 5-year longitudinal study, further found that MIND diet adherence was associated with larger hippocampal volumes and demonstrated that circulating polyphenol, omega-3, and B-vitamin levels were related to both reduced neuroinflammation and improved cognition (19). On the other hand, another 3-year longitudinal study in healthy older adults found no associations between MIND diet adherence and systemic inflammation markers nor links with cognition (20). These inconsistent findings emphasize the need to examine the relationship between MIND diet and inflammation in more detail. Moreover, the role of IB permeability in the relation between MIND diet adherence and systemic inflammation has not been investigated yet in older adults. Critically, low-grade systemic inflammation has been implicated in age-related cognitive decline (21), as peripheral inflammation can reach the central nervous system and promote neuroinflammation (22).

Neuroinflammation refers to the activation of central nervous system immune cells, primarily microglia and astrocytes. Similar to peripheral inflammation, the ageing brain can exhibit a prolonged low-grade state of neuroinflammation (23). Studies using proton magnetic resonance spectroscopy (^1^H-MRS) in older adults have revealed elevated levels of metabolites (indirectly) associated with neuroinflammation (myo-inositol, creatine, and choline (24)) in various brain regions, including the dorsolateral prefrontal cortex (dlPFC) and hippocampus (25,26). These specific regions are known for their vulnerability to both ageing and neuroinflammation (27,28). Importantly, neuroinflammation plays a crucial role in ageing-related cognitive decline and AD (29,30), with increased brain neuroinflammation-associated metabolites inversely relating to cognitive performance in aged individuals (25,31). Although neuroinflammation can originate from numerous (complex) processes, it may also result directly from systemic inflammation (32). Peripherally-produced cytokines can access the brain via neural pathways (the vagus nerve) or vascular routes (crossing the blood-brain barrier) (33). Furthermore, these cytokines can disrupt blood-brain barrier (BBB) integrity (34), facilitating the infiltration of circulating immune cells and plasma proteins into brain tissue (35). Indeed, middle-aged adults with elevated CRP levels demonstrated increased cerebral myo-inositol levels measured with ^1^H-MRS (36). Moreover, a longitudinal study using structural neuroimaging showed that low-grade systemic inflammation (measured with the INFLA-score) was associated with reduced brain volumes, especially in the cortex (37).

Evidence linking MIND diet adherence to neuroinflammation is scarce. One post-mortem study assessed hippocampal microglia in individuals without dementia and concluded that a higher MIND diet adherence was associated with fewer reactive microglia (38). In addition, an RCT showed that Mediterranean-type diets modulated expression of neuroinflammation-related genes (39). However, in vivo human evidence (using neuroimaging techniques) linking Mediterranean and MIND diet adherence to neuroinflammation is currently lacking. Moreover, no studies have assessed the relationship between MIND diet adherence and the peripheral-to-central inflammatory link.

Therefore, we aimed to explore the relation between MIND diet adherence (Dutch version: MIND-NL) (8), systemic inflammation (based on CRP and WBC parameters), neuroinflammation (based on myo-inositol, choline, and creatine measured with ^1^H-MRS within the dlPFC), and cognitive functioning cross-sectionally. We also specifically assessed whether MIND diet adherence moderates the link between peripheral and central inflammation, and ultimately cognitive functioning. Additionally, we assessed to what extent IB permeability could explain systemic inflammation and potential further relations with neuroinflammation and cognition.

## Methods

### Study design

We used baseline data from an RCT study, named HELI, and performed a cross-sectional analysis. The HELI study is a 26-week multicenter randomized controlled multidomain lifestyle intervention trial in older adults at risk of cognitive decline. Details about the study intervention groups, outcomes, and procedures can be found in the study design paper (40). The study protocol was approved by the Medical Research Ethics Committee Oost-Nederland (ToetsingOnline filenumber NL78263.091.21, ClinicalTrials.gov ID NCT05777863). Each participant provided written consent.

### Participants

Participants were enrolled between May 2022 and October 2023. Individuals had to be (1) between 60-75 years old at moment of signing written informed consent, (2) fluent in the Dutch language, and (3) score ≥2 points on the self-reported ‘Modifiable cardiovascular risk factor scale’ (**Supplementary Table 1**). This risk factor scale (adapted from CAIDE (41)) was used as a tool to indicate increased risk for cognitive decline, based on lifestyle-modifiable risk factors. Individuals were excluded when they (1) concurrently participated in other intervention trials, (2) had no access to technological infrastructure (i.e., computers, smartphone, mobile applications, internet access, etc.), (3) were cognitively impaired as determined by a Telephone Interview for Cognitive Status (TICS-M1) score <23 points, (4) had a clinical diagnosis of a cerebrovascular event, neurological disease, current malignant disease, current psychiatric disorder, moderate to severe cardiovascular disease, revascularization surgery in the last 12 months, inflammatory bowel disease, visual impairment (which is uncorrectable with visual aids), and hearing or communicative impairment (which is uncorrectable with hearing aids), or (5) had MRI contradictions. A total of 102 participants were included in the intervention. Participants were excluded from the current analysis in case of missing data, when identified as an outlier, or in case of insufficient data quality. Outlier criteria and/or data quality assessment steps and resulting exclusions of participants are explained per outcome, when applicable. In the end, 88 participants were included in the current analysis.

### MIND-NL diet adherence

Participants filled in a short validated Dutch Food Frequency Questionnaire (MIND-NL-Eetscore-FFQ), with the previous month as a reference. This condensed FFQ contains 72 food items and 41 questions and evaluates both compliance with the 2015 Dutch dietary guidelines and the MIND-NL diet. Questions are structured chronologically, starting with breakfast and advancing to late evening. Intake estimates were derived using standard portion sizes and commonly used household measures. Per food item, mean daily consumption (in grams) was determined by multiplying consumption frequency by portion size. The MIND-NL-Eetscore-FFQ was implemented through a web-based assessment tool (www.eetscore.nl).

Here, we used the MIND-NL diet adherence score, which is adapted to the Dutch context and serving sizes, and captures dietary consumption of 15 food groups, comparable to the original MIND diet (8). Nine of the 15 food groups are recommended: green leafy vegetables, other vegetables, berries and strawberries, whole grains, nuts, fish, poultry, beans and legumes, and olive oil. Six of the 15 food groups are discouraged: full-fat cheese, butter and sticky margarines, red and processed meat, take out meals, fried foods and snacks, cookies, pastries and sweets, and wine and other alcoholic beverages. Each food group received a score of 0, 0.5, or 1.0, with higher scores meaning a better adherence to the food group (higher consumption in case of recommended food group, lower consumption in case of discouraged food group). The MIND-NL score combines the individual food group scores into a total score ranging from 0 to 15, similar to the original MIND diet. Higher scores indicate better adherence to the MIND-NL diet. Cut-off scores per food group can be found in **Supplementary Table 2.** We refer to the paper of Beers et al. (8) for an extensive description of the MIND-NL diet components and scoring system.

### Blood measurements

Blood was drawn in a fasted state in the morning. On the day before the study visit, participants consumed a standardized meal (610 kJ energy, 1.70 g fat, 23.9 g carbohydrates, 5.30 g fiber, and 6.00 g protein per 100 g) and fasted from 20:00. Blood was drawn via a finger prick, and intravenously from the arm.

#### Systemic inflammation markers

We measured hs-CRP concentrations and WBC counts to assess systemic inflammation levels. These markers were determined directly in capillary blood obtained via fingerstick. For hs-CRP measurement, 20 µL of blood was collected in a capillary tube, transferred into a cuvette with buffer solution, and analyzed using the QuikRead Go analyzer (Orion Diagnostics, Ghodbunder, India) following the manufacturer’s instructions (measurement range: 0.5–200 mg/L). For total and differential WBC counts (neutrophils, lymphocytes, monocytes, eosinophils, and basophils), 10 µL of blood was collected in a microcuvette and analyzed using the HemoCue® WBC DIFF (HemoCue AB, Ängelholm, Sweden) according to the manufacturer’s instructions (measurement range: 0.3–30.0×10L/L).

Given our focus on chronic low-grade systemic inflammation, participants with signs of acute infection were excluded from the analysis. Specifically, we excluded participants with CRP values >22 mg/L. In healthy individuals, CRP values between 3 and 10 mg/L generally represent low-grade systemic inflammation, whereas values >10 mg/L may indicate acute infection (13). However, among individuals with obesity, CRP values ≤22 mg/L can still reflect low-grade systemic inflammation without acute infection (42). Given that our sample consisted primarily of overweight and obese participants, we retained participants with CRP values between 10 and 22 mg/L. For these participants, as well as for those with outlying values in other inflammatory markers (>1.5× IQR), we reviewed the general wellbeing questionnaire completed during the study visit to identify potential underlying infections. Participants were included only if they reported no acute illnesses (e.g., cold, flu) or other potential inflammatory conditions. In total, 1 participant reporting an active infection was excluded from the analysis.

#### Intestinal barrier permeability markers

Three biomarkers indicating bacterial translocation and/or IB permeability were measured in serum: lipopolysaccharide (LPS), LPS-binding protein (LBP), and zonulin (43). ELISAs were used to quantify LBP and zonulin in 25 µL of serum each. LBP was measured using the HK315-02 assay kit (Hycult Biotech, Uden, Netherlands), and zonulin was measured using the K5601 assay kit (Immundiagnostik AG, Bensheim, Germany), following manufacturers’ instructions. LPS was measured in serum with a chromogenic assay from Nodia (associates Cap. Cod Inc.). The assay was performed in a Pyros Kinetix Flex tube reader (pKFlex) (Nodia, Cap. Cod Inc.). Pyrochrome lysate was reconstituted with 3.4 ml Pyrochrome reconstitution buffer (C1500-5) and for the calibration curve Limulus Amebocyte lysate control standard with an endotoxin concentration of 0.5 µg/vial (CSEE0005-1) was used. The samples were diluted with LPS free MQ and incubated for 15 min in a water bath at 70 °C. After 1 hour at 4 °C the samples were placed for 15 min at room temperature and directly measured on the pKflex. 200 µl treated sample or standard was added in a pKFlex glass tube, 50 µl Pyrochrome was added, and the mixture was shortly stirred and placed in the pKFlex. Using the Pyrosexpress 21 CFR Part 11 compliant software the endotoxin concentration was calculated. All samples were analyzed in duplicate. IB permeability markers were only analyzed in participants that completed the intervention study. From the *N*=88 participants that were included in the current study, IB permeability lab analysis was not performed in *n*=17 participants due to intervention drop-out reasons. Therefore, the additional analyses with intestinal permeability markers were performed in *n*=71 participants.

### Neuroinflammation using ^1^H-MRS metabolites

To assess neuroinflammation, we measured intracranial levels of neuroinflammation-associated metabolites using ^1^H-MRS. ^1^H-MRS data were initially acquired in the left dlPFC and hippocampus. However, due to general insufficient spectral quality, hippocampal data could not be used for analysis. For this reason, we only used dlPFC spectral data for the quantification of neuroinflammation metabolites. We used a Siemens Healthineers Magnetom Skyra 3 Tesla MRI scanner with a 32-channel MR head coil. A high-resolution T1-weighted whole-brain anatomical scan was acquired using a magnetization-prepared 2 rapid gradient echo (MP2RAGE) sequence (TR/TE = 6000/2.34 ms; voxel size = 1.0 × 1.0 × 1.0 mm3; FoV = 256 mm; flip angle = 6°; 176 sagittal slices; interleaved slice acquisition; 1.0 mm slice thickness). ^1^H-MRS metabolite spectra in a 20 x 20 x 20 mm isotropic voxel were acquired with a Point RESolved Spectroscopy (PRESS) sequence (TR/TE = 2000/35 ms; averages = 64; flip angle = 90°) using chemically selective water suppression (CHESS) (44). In addition, water-unsuppressed reference data were acquired (TR/TE = 2000/35 ms; averages = 16; flip angle = 90°). To minimize field inhomogeneities within the isotropic voxel, advanced shimming was used. Voxel location in the left dlPFC was based on a fast T1 weighted scan (TR/TE = 6.3/3.2 ms; voxel size = 1.0 × 1.0 × 1.0mm; FoV = 256 mm; flip angle = 11°) and was performed across participants manually based on a reference image (**Figure 1a**). To maintain data quality, participant-specific voxel adjustments were applied to account for individual anatomical variations in the cortical region, minimizing cerebrospinal fluid (CSF) inclusion and excluding non-brain tissue.

**Figure 1.**
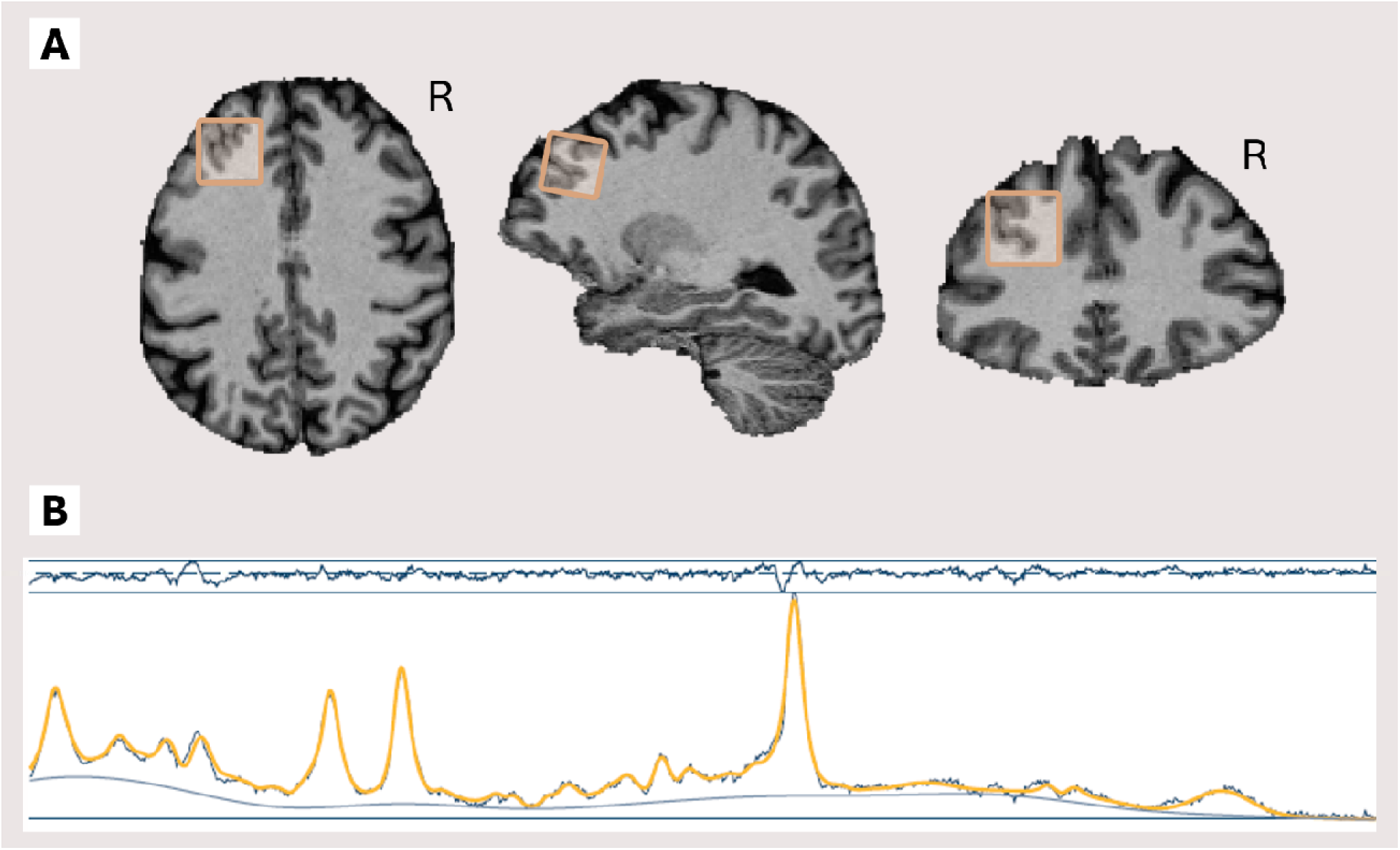
^1^H-MRS example voxel and spectrum. Transversal, sagittal and coronal image of voxel location in the left dlPFC retrieved from example participant (**A)**. Fitted spectrum of example participant (**B**). The oscillating dark blue line on top represents the residuals. The yellow line represents the fitted spectrum (i.e. the modelled spectrum that is used for quantification of metabolites) and the blue line represents the observed spectrum (aligned and averaged). The blue line below the observed and fitted spectra represents a regressor for chemical shift drift.

We were interested in three brain metabolites to quantify neuroinflammation: myo-inositol, total choline, and total creatine (25). Osprey 2.4.0 (45), an open-source software implemented in MATLAB R2024a (Mathworks Inc.; https://nl.mathworks.com/products/matlab.html), was used to process ^1^H-MRS data. Osprey provides automated and standardized preprocessing, linear-combination modeling, tissue correction, and quantification of metabolites. Acquired data (metabolite, lineshape reference, and anatomical) were preprocessed, generating averaged spectra. Osprey applies automated eddy-current corrections (46) based on water-unsuppressed reference data. The preprocessed, averaged spectra were fitted using Osprey’s linear combination model with default settings. Metabolite spectra were fitted across a frequency range of 0.5 to 4.0 ppm with a knot spacing of 0.4 ppm. Water data was fitted in the range of 2.0 to 7.4 ppm. Spectral data were coregistered to the participant’s T1-weighted anatomical scan (MP2RAGE). To derive fractional tissue volumes within the voxel location, tissue segmentation was performed on the anatomical data using Statistical Parametric Mapping 12 (SPM12; https://www.fil.ion.ucl.ac.uk/spm/software/spm12/). To quantify molar concentration estimates of myo-inositol, total choline, and total creatine, tissue (relaxation time) corrected and water-scaled metabolite measures were calculated (47,48). Metabolite concentrations were expressed in mol/kg. See **Figure 1b** for a fitted spectrum of an example participant.

To evaluate spectral quality, we manually inspected pre-aligned, post-aligned, averaged, and fitted spectra. Specifically, we assessed the following quality metrics: absence of lipid peaks (1-2 ppm), adequate water suppression (3-4 ppm), presence of peaks for metabolites of interest (creatine at 3.03 ppm, choline at 3.20 ppm, and myo-inositol at 3.5 and 4.06 ppm), absence of chemical shift drift indicating movement artifacts, and adequate shimming. In addition, we assessed Osprey’s reported data quality measures including creatine signal-to-noise (SNR) ratio and full width at half maximum (FWHM) values. Participants were excluded from analysis in case of insufficient spectral quality. This led to the exclusion of *n*=10 participants from our dataset. See **Supplementary Table 3** for a summary of data quality measures and tissue fractions in our dataset.

### Cognitive functioning

To measure cognitive functioning, participants performed a neuropsychological test battery (NTB). The NTB was focused on cognitive domains predominantly affected in ageing and consisted of: the Digit Span Test (DST) to measure working memory [58], the Digit Symbol Substitution Test (DSST) to assess processing speed [58], the Rey Auditory Verbal Learning Test (RAVLT) to evaluate episodic memory [59], and the Trail Making Test (TMT) [60] and Verbal Fluency Test (VFT) [61] to assess executive functioning. We used the following outcomes: DST total score (forward+backward), DSST total score, RAVLT delayed recall score, TMT-B/TMT-A ratio score, and VFT total score. Individual cognitive tests scores were Z-scored and combined into a composite cognition Z-score with higher values indicating a better cognitive performance, attributing an equal weight to each neuropsychological test. Additionally, we conducted the Montreal Cognitive Assessment (MoCA) as a descriptive measure of cognitive function.

### Data analysis

#### Combination scores

Since we measured multiple biomarkers involved in the same biological processes, we decided to combine these markers in one score. To create these combination scores, we linearly combined relevant markers for each biological outcome (i.e. systemic inflammation, neuroinflammation, and IB permeability) using principal component analyses (PCAs). Individual biomarker data were centered and scaled before processed in the PCA. The first principal component (PC1) scores per participant were retrieved for each biological outcome and used in further analyses.

For the systemic inflammation combination score, we selected blood CRP levels, WBC counts, and neutrophil-to-lymphocyte ratio (NLR), based on the INFLA-score (14). For the neuroinflammation combination score, we selected myo-inositol, total choline, and total creatine levels in the dlPFC (25). For the IB permeability combination score, we selected blood LBP, zonulin, and LPS levels (43). In case individual biomarkers loaded negatively on PC1, PC1 scores were reversed, meaning that a higher PC1 score represents higher presence of the selected biomarkers, thus higher systemic inflammation, neuroinflammation, or IB permeability.

#### Statistical analysis

Statistical analyses were performed in R version 4.3.2. Pairwise correlations between individual outcomes (MIND diet adherence, systemic inflammation, neuroinflammation, cognition) were assessed using Spearman correlation analyses. Based on pairwise correlation results, we used multiple linear regression models to predict 1) neuroinflammation and 2) cognition with MIND diet adherence and systemic inflammation as independent variables. To identify a potential moderating role of MIND diet adherence, each linear model was analyzed with and without an interaction between MIND diet adherence and systemic inflammation. In case of a significant interaction effect, post-hoc simple slopes and Johnson-Neyman analyses were performed. Linear models were conducted using the lm function from the stats package in R.

Based on multiple linear regression model results, a mediation analysis was selected as next step, to identify potential mediating roles within the framework of MIND diet adherence, systemic inflammation, neuroinflammation, and cognition. Mediation analyses were conducted using structural equation modeling (SEM) via the lavaan package in R (49). The mediation model specified regression paths from the predictor to the mediator (path a), from the mediator to the outcome controlling for the predictor (path b), and the direct effect from predictor to outcome controlling for the mediator (path c’). The indirect effect was computed as the product of paths a and b. Bootstrap confidence intervals were calculated with 5000 resamples.

Within all models, sex and age were used as covariates. Models predicting neuroinflammation included relResA as covariate, a ^1^H-MRS spectral quality measure. Models predicting cognitive functioning included education levels as covariate. The normality of residuals was assessed using Q-Q plots, and homoscedasticity (equal variance) was evaluated by plotting residuals against the fitted values. P-values ≤ 0.05 were considered statistically significant.

To assess the link between IB permeability and systemic inflammation, and to examine further relations with neuroinflammation and cognition, we used similar statistical models and analysis steps as described above. We started with Spearman correlations to explore the relation between IB permeability and the other outcomes. Based on the observed correlations, we employed linear- and mediation models. Full model statistics of all used models can be found in **Supplementary Tables 4, 5, and 6**.

## Results

### Population characteristics

Within our study population (*N*=88), the mean age was 66.4 ± 4.4 years, 67% of the participants was female and the mean BMI was 29.9 ± 5.2 kg/m^2^ (**Table 1**). The mean MIND diet adherence score was 8.6 ± 2.0 and 30% of the participants had a CRP above 3 mg/L (**Table 2**).

**Table 1.**
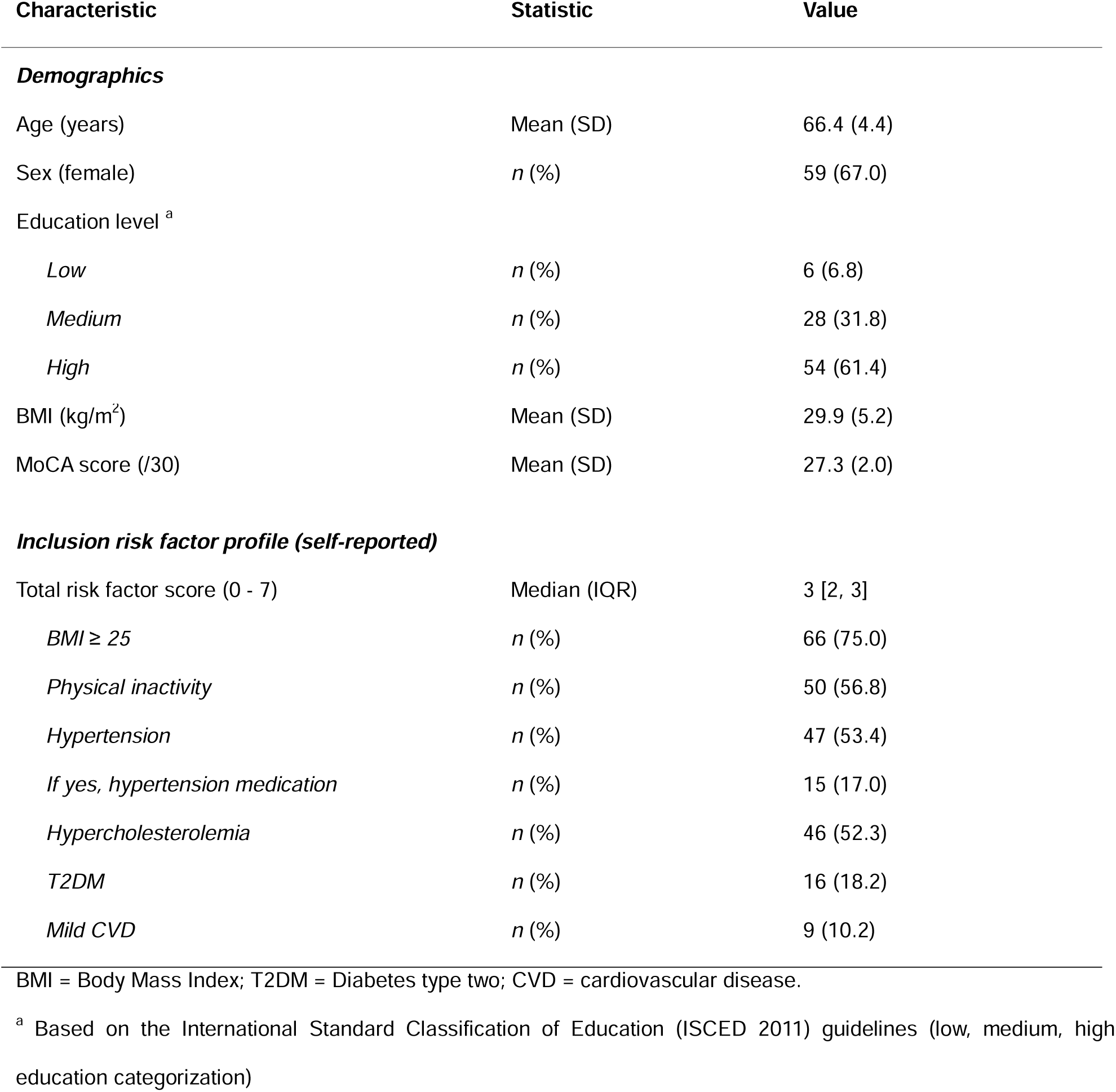
Population characteristics (*N*=88)

| Characteristic | Statistic | Value |
| --- | --- | --- |
| **Demographics** |  |  |
| Age (years) | Mean (SD) | 66.4 (4.4) |
| Sex (female) | *n* (%) | 59 (67.0) |
| Education level <sup>a</sup> |  |  |
| *Low* | *n* (%) | 6 (6.8) |
| *Medium* | *n* (%) | 28 (31.8) |
| *High* | *n* (%) | 54 (61.4) |
| BMI (kg/m<sup>2</sup>) | Mean (SD) | 29.9 (5.2) |
| MoCA score (/30) | Mean (SD) | 27.3 (2.0) |
| **Inclusion risk factor profile (self-reported)** |  |  |
| Total risk factor score (0 - 7) | Median (IQR) | 3 [2, 3] |
| *BMI ≥ 25* | *n* (%) | 66 (75.0) |
| *Physical inactivity* | *n* (%) | 50 (56.8) |
| *Hypertension* | *n* (%) | 47 (53.4) |
| *If yes, hypertension medication* | *n* (%) | 15 (17.0) |
| *Hypercholesterolemia* | *n* (%) | 46 (52.3) |
| *T2DM* | *n* (%) | 16 (18.2) |
| *Mild CVD* | *n* (%) | 9 (10.2) |

BMI = Body Mass Index; T2DM = Diabetes type two; CVD = cardiovascular disease.
<sup>a</sup> Based on the International Standard Classification of Education (ISCED 2011) guidelines (low, medium, high education categorization)

**Table 2.** Descriptive summary of study outcomes (*N*=88)

| Study outcome | Statistic | Value |
| --- | --- | --- |
| MIND-NL diet adherence score (/15) | Median (IQR) | 8.75 [7, 10] |
| Systemic inflammation |  |  |
| *CRP (mg/L)* | Median (IQR) | 1.5 [1.2, 3.9] |
| *0-3 mg/L, n (%)* | *n* (%) | 62 (70.5) |
| *3-10 mg/L, n (%)* | *n* (%) | 22 (25) |
| *10-22 mg/L, n (%)* | *n (%)* | 4 (4.5) |
| *WBC (counts×10<sup>9</sup>/L)* | Mean (SD) | 6.3 (1.8) |
| *NLR* | Mean (SD) | 1.4 (0.5) |
| Neuroinflammation |  |  |
| *dIPFC myo-inositol (mol/kg)* | Mean (SD) | 10.1 (1.3) |
| *dIPFC total choline (mol/kg)* | Mean (SD) | 2.8 (0.4) |
| *dIPFC total creatin (mol/kg)* | Mean (SD) | 12.8 (1.1) |
| Cognitive test scores |  |  |
| *DST (forward + backward total score) (/28)* | Mean (SD) | 15.6 (3.4) |
| *DSST (total score) (/90)* | Mean (SD) | 52.8 (10.0) |
| *RAVLT (delayed recall score) (/15)* | Mean (SD) | 7.9 (3.1) |
| *TMT (TMT-B/TMT-A score)* | Mean (SD) | 2.2 (0.7) |
| *VFT (total score)* | Mean (SD) | 26.5 (5.4) |
| Intestinal barrier permeability <sup>a</sup> |  |  |
| *LBP (ug/mL)* | Mean (SD) | 13.6 (4.0) |
| *Zonulin (ng/mL)* | Mean (SD) | 38.6 (17.2) |
| *LPS (EU/mL)* | Median (IQR) | 1.01 [0.33, 2.72] |

CRP = C-reactive protein; WBC = white blood cell; NLR = neutrophil-to-lymphocyte ratio; dIPFC = dorsolateral prefrontal cortex; DST = digit span test; DSST = digit symbol substitution test; RAVLT = Rey auditory verbal learning test; TMT = trail making test; VFT = verbal fluency test; LBP = lipopolysaccharide binding protein; LPS = lipopolysaccharide.
<sup>a</sup> For intestinal barrier permeability markers, *n*=71

#### Combination scores based on principal component analysis

To integrate individual biomarkers reflecting the same biological process into composite scores, we performed PCAs and extracted PC1 as a linearly weighted summary measure. PC1 per biological outcome explained 57.8% (systemic inflammation), 65.7% (neuroinflammation), and 46.8% (IB permeability) of the total variance. PCA biplots and PC1 loadings per individual biomarker can be found in **Supplementary Figure 1** for each biological outcome.

### No direct relation between MIND diet adherence, systemic inflammation, neuroinflammation, and cognitive functioning

To explore the relation between our main outcomes of interest (MIND diet adherence, systemic inflammation, neuroinflammation, and cognitive functioning), we employed Spearman correlation analyses for each pair of outcomes. No significant correlations were found between MIND diet adherence, systemic inflammation, neuroinflammation, and cognitive functioning (**Supplementary Figure 2a-f**). A correlation heatmap containing all components of the combination scores (e.g. all individual biomarker and cognitive scores) and relevant other variables (e.g. covariates) can be found in **Supplementary Figure 3**.

Beyond total MIND diet adherence, we examined adherence to (consuming) recommended versus (limiting) discouraged food groups separately. MIND diet adherence to discouraged food groups was significantly correlated with systemic inflammation (*Spearman’s rho*=-0.28; *p*=0.009), showing that lower consumption of discouraged food groups was associated with lower systemic inflammation (**Supplementary Figure 3 and 4)**.

### MIND diet adherence moderation of the relation between systemic inflammation and neuroinflammation

As a next step, we employed linear models to predict neuroinflammation with MIND diet adherence and systemic inflammation. Although MIND diet adherence (β=0.00, *p*=0.97) and systemic inflammation (β=0.12, *p*=0.30) independently did not predict neuroinflammation (**Supplementary Table 4, model 1a)**, the interaction between MIND diet adherence and systemic inflammation significantly predicted neuroinflammation (β=-0.11, *p*=0.04), when added to the model (**Supplementary Table 4, model 1b).** Specifically, systemic inflammation significantly predicted neuroinflammation when MIND diet adherence was lower (-1SD; β=0.37, *p*=0.03), and not when MIND diet adherence was average (mean; β=0.14, *p*=0.24) or higher (+1SD; β=-0.09, *p*=0.55) (**Figure 2a)**. To determine the threshold MIND diet adherence score, we conducted a Johnson-Neyman analysis. The relation between systemic inflammation and neuroinflammation was significant for MIND diet adherence scores lower than 7.4 (**Figure 2b)**.

**Figure 2.**
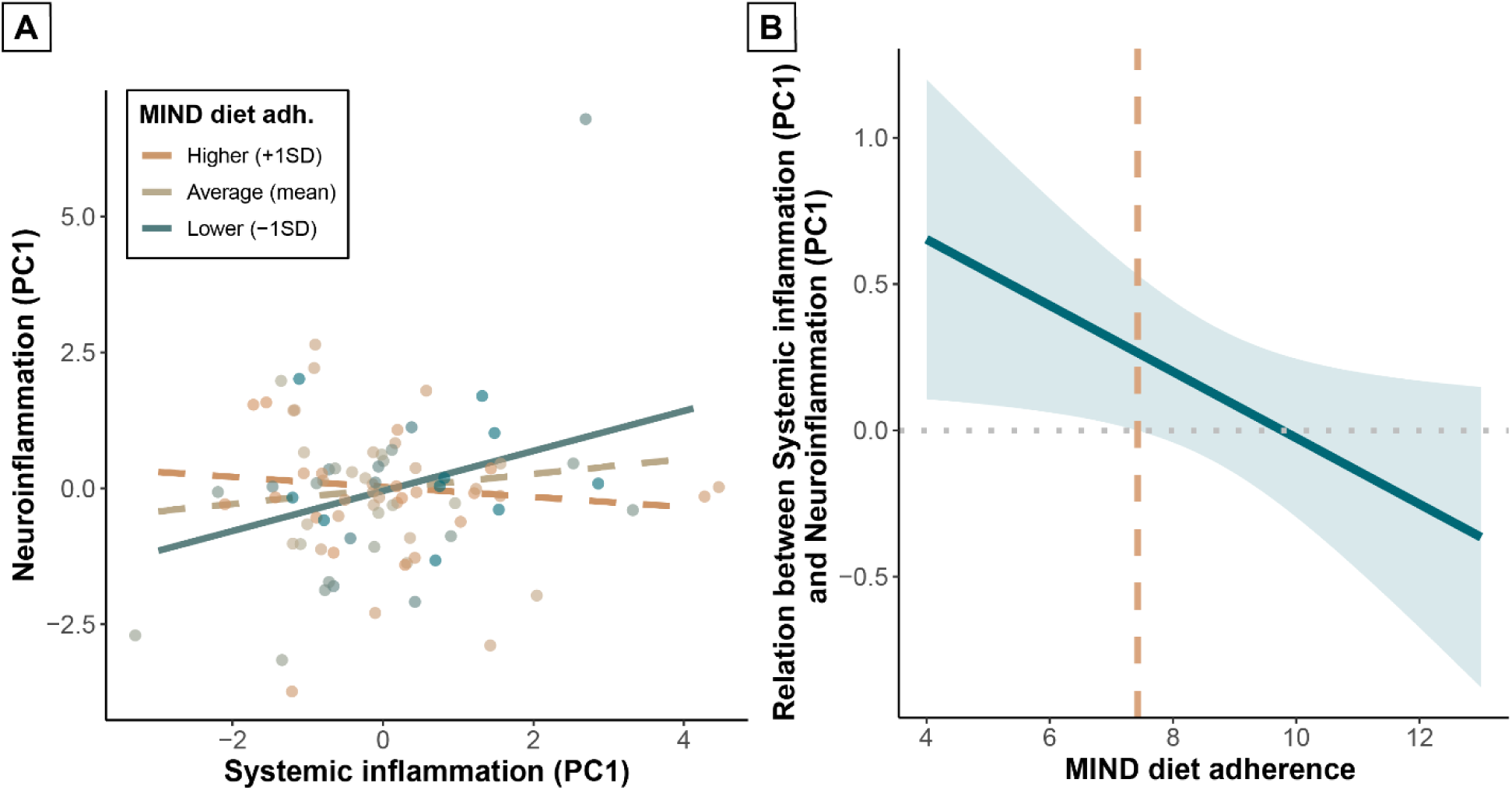
Interaction between MIND diet adherence and systemic inflammation on neuroinflammation (N=88). Interaction plot (**A**) showing the relation between systemic inflammation and neuroinflammation for lower (-1SD), average (mean), and higher (+1SD) MIND diet adherence. Solid lines represent significant slopes; dashed lines represent non-significant slopes. Dots represent individual participants; color grades indicate MIND diet adherence subgroups. Johnson-Neyman plot (**B**) showing the relation between systemic inflammation and neuroinflammation for MIND diet adherence scores. Trendline and confidence interval are displayed in blue. Orange dashed line represents the threshold MIND diet adherence score for a significant relation. Linear model was corrected for age, sex, and relResA/spectral quality.

### MIND diet adherence moderation of the relation between systemic inflammation and cognitive functioning

Similar to neuroinflammation, we found that MIND diet adherence (β=0.01, *p*=0.75) and systemic inflammation (β=-0.02, *p*=0.55) did not independently predict cognition (**Supplementary Table 4, model 2a)**. However, again, we did observe a significant interaction between MIND diet adherence and systemic inflammation when adding the interaction term to the model (β=0.044, *p*=0.02) (**Supplementary Table 4, model 2b)**. Systemic inflammation significantly predicted cognition when MIND diet adherence was lower (-1SD; β=-0.12, *p*=0.03), and not when MIND diet adherence was average (mean; β=-0.03, *p*=0.47) or higher (+1SD; β=0.06, *p*=0.24) (**Figure 3a)**. The relation between systemic inflammation and cognition was significant for MIND diet adherence scores lower than 7.0, based on a Johnson-Neyman analysis (**Figure 3b)**.

**Figure 3.**
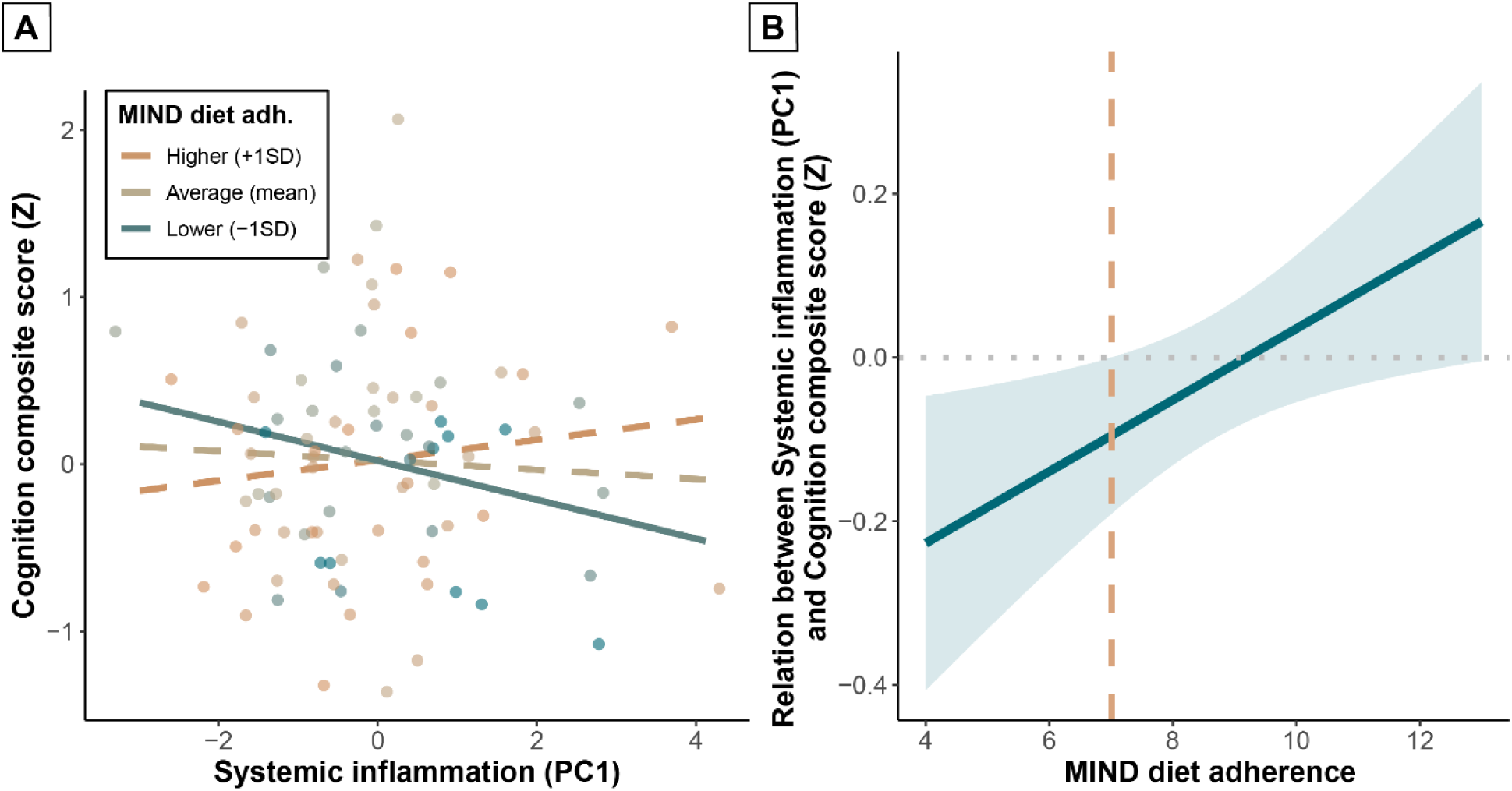
Interaction between MIND diet adherence and systemic inflammation on cognition (N=88). Interaction plot (**A**) showing the relation between systemic inflammation and cognition for lower (-1SD), average (mean), and higher (+1SD) MIND diet adherence. Solid lines represent significant slopes; dashed lines represent non-significant slopes. Dots represent individual participants; color grades indicate MIND diet adherence subgroups. Johnson-Neyman plot (**B**) showing the relation between systemic inflammation and cognition for MIND diet adherence scores. Trendline and confidence interval are displayed in blue. Orange dashed line represents the threshold MIND diet adherence score for a significant relation. Linear model was corrected for age, sex, and education.

To assess whether the relationship between the MIND diet adherence-systemic inflammation interaction and cognition could be fully explained by neuroinflammation, we employed a mediation analysis. This analysis showed that neuroinflammation did not significantly mediate the relation between the MIND diet adherence-systemic inflammation interaction and cognition (β=0.003 [-0.005, 0.019], *p*=0.58) (**Supplementary Figure 5**). Full mediation model statistics can be found in **Supplementary Table 5**.

### MIND diet adherence moderation of the relation between intestinal barrier permeability and neuroinflammation

To examine the role of IB permeability within the framework of MIND diet adherence, systemic inflammation, neuroinflammation, and cognition, we conducted Spearman correlation analyses to test the relation of IB permeability with these outcomes as a first step. IB permeability significantly correlated with systemic inflammation (*Spearman’s rho*=0.40, *p*<0.001) (**Figure 4a**), but not with MIND diet adherence, and neuroinflammation (**Supplementary Figure 2g-h**). A trend negative correlation between IB permeability and cognition was found (*Spearman’s rho*=-0.21, *p*=0.08) (**Supplementary Figure 2i**). As a next step, we employed linear models to predict neuroinflammation with MIND diet adherence and IB permeability instead of systemic inflammation. MIND diet adherence (β=-0.01, *p*=0.91) and IB permeability (β=0.15, *p*=0.34) did not independently predict neuroinflammation (**Supplementary Table 4, model 3a)**, but, again, adding the interaction between MIND diet adherence and IB permeability to the model showed a significant prediction effect (β=-0.17, *p*=0.05) (**Supplementary Table 4, model 3b)**. Specifically, IB permeability significantly predicted neuroinflammation when MIND diet adherence was lower (-1SD; β=0.59, *p*=0.03), but not when MIND diet adherence was average (mean; β=0.25, *p*=0.13) or higher (+1SD; β=-0.10, *p*=0.62) (**Figure 4b)**. The relation between IB permeability and neuroinflammation was significant for MIND diet adherence scores lower than 7.7, based on a Johnson-Neyman analysis (**Figure 4c)**.

**Figure 4.**
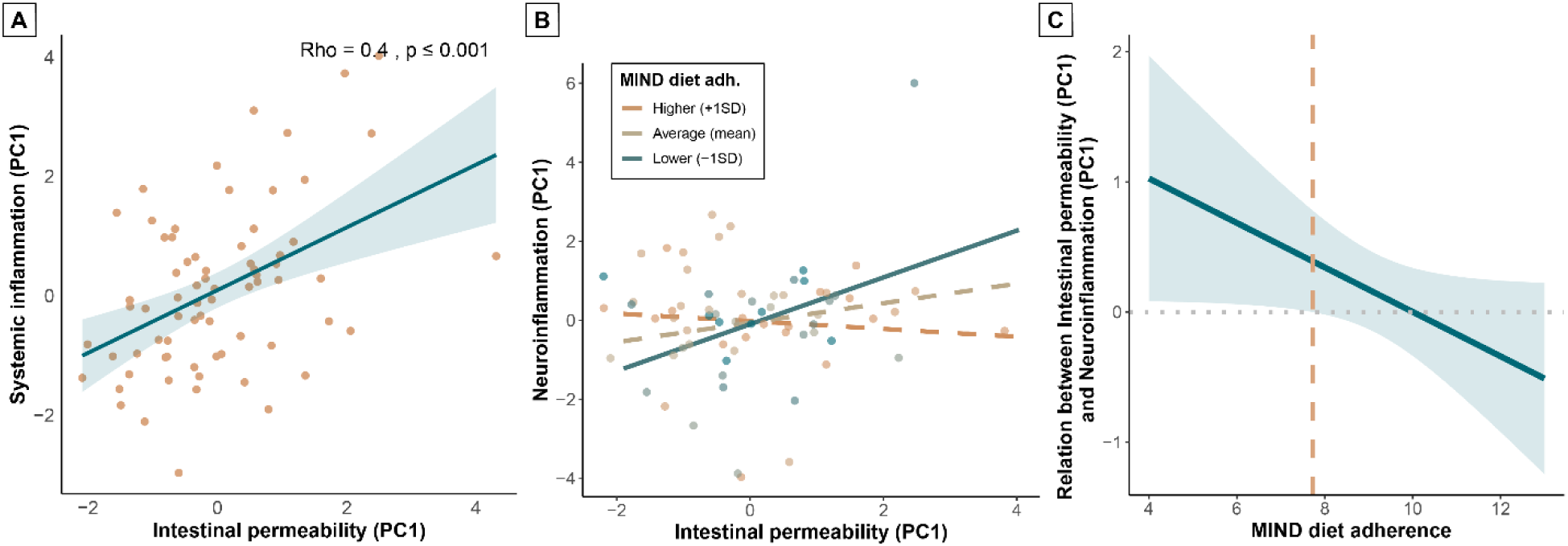
Role of intestinal barrier permeability in the relation between MIND diet adherence, systemic inflammation and neuroinflammation (n=71). Spearman correlation scatterplot (**A**) showing correlation between intestinal barrier permeability and systemic inflammation. Orange dots represent individual participants; trendline and confidence interval are displayed in blue. Spearman’s rho and p-value are reported top right. Data was corrected for age and sex; residuals were used for correlation analyses. Interaction plot (**B**) showing the relation between intestinal barrier permeability and neuroinflammation for lower (-1SD), average (mean), and higher (+1SD) MIND diet adherence. Solid lines represent significant slopes; dashed lines represent non-significant slopes. Dots represent individual participants; color grades indicate MIND diet adherence subgroups. Johnson-Neyman plot (**C**) showing the relation between intestinal barrier permeability and neuroinflammation for MIND diet adherence scores. Trendline and confidence interval are displayed in blue. Orange dashed line represents the threshold MIND diet adherence score for a significant relation. Linear model was corrected for age, sex, and relResA/spectral quality.

As a final step, to assess whether the relationship between the IB permeability and neuroinflammation within the group with lower MIND diet adherence could be explained by systemic inflammation, we conducted a mediation analysis within the individuals with lower MIND diet adherence (*n*=35, adherence score ≤8.75) and the individuals with higher MIND diet adherence (*n*=36, adherence score >8.75) based on a median split. We found that systemic inflammation mediated the relation between the IB permeability and neuroinflammation within the low MIND diet adherence group (β=0.427 [0.072; 0.891], *p*=0.04), but not within the high MIND diet adherence group (β=-0.064 [-0.298; 0.081], *p*=0.51) (**Figure 5**). Full mediation model statistics can be found in **Supplementary Table 6**.

**Figure 5.**
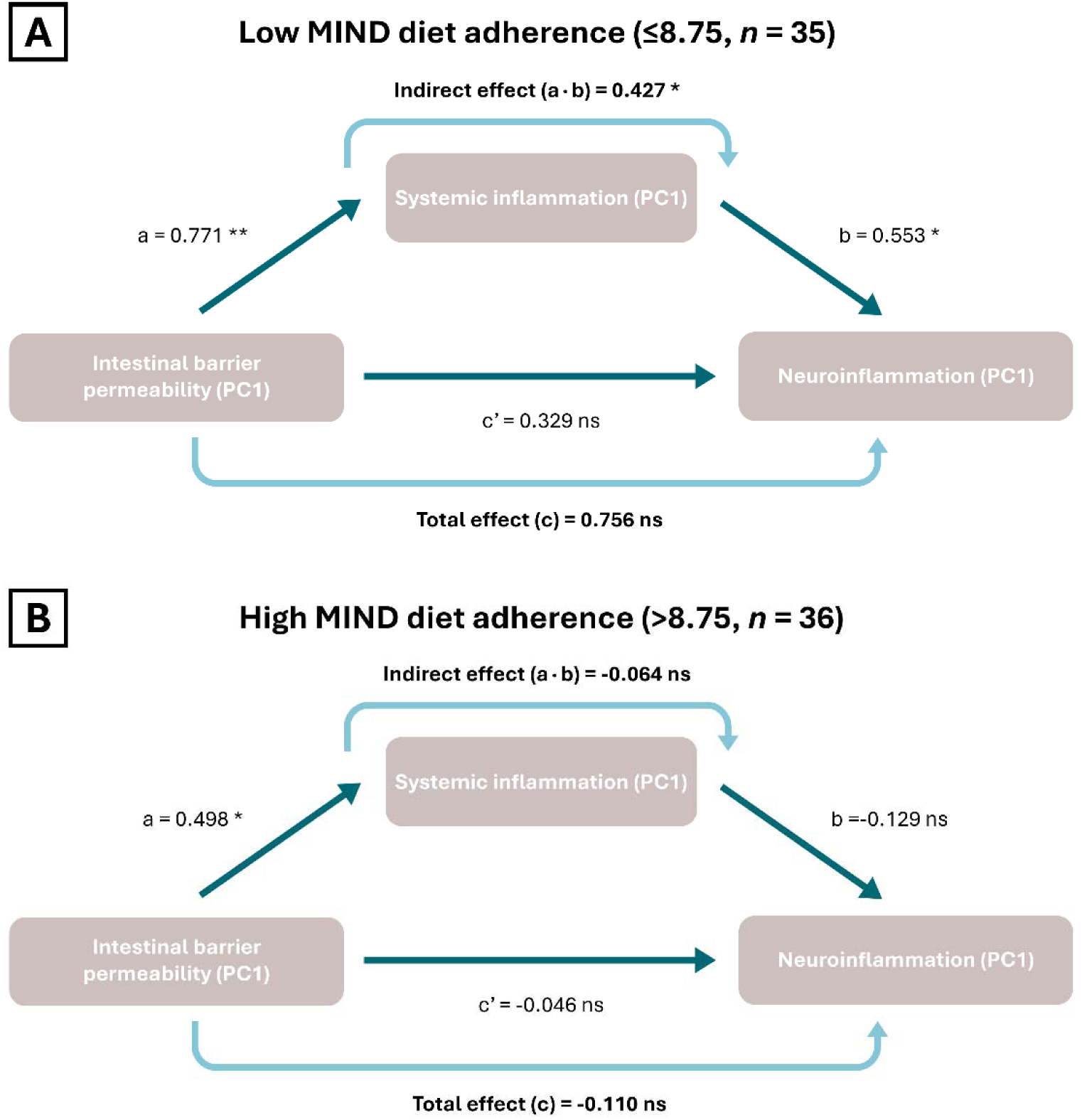
Mediation analysis diagram for intestinal barrier permeability, systemic inflammation, and neuroinflammation in low (n=35) and high (n=36) MIND diet adherence group. Diagram showing the mediating role of systemic inflammation in the relation between intestinal barrier permeability and neuroinflammation, within low MIND diet adherence group (score ≤8.75) and high MIND diet adherence group (score >8.75) based on median split. Significance is indicated with asterisks (*** = 0.001, ** = 0.01, * = 0.05, ns = non-significant).

## Discussion

The goal of this study was to explore the cross-sectional associations between MIND diet adherence, systemic inflammation, neuroinflammation, cognitive functioning, and IB permeability in older adults. Our results demonstrate that MIND diet adherence moderates the relation between systemic inflammation and neuroinflammation as well as cognitive functioning in older adults. Specifically, the association between systemic inflammation and both neuroinflammation and cognitive functioning was only evident in individuals with lower MIND diet adherence. In addition, we showed that in individuals with lower MIND diet adherence, the relation between IB permeability and neuroinflammation is mediated by systemic inflammation. The moderating effect of MIND diet adherence on these peripheral-central relations is robustly observed, with significant associations present below threshold adherence scores between 7 and 8. Consequently, we demonstrated novel associations between MIND diet adherence, IB permeability, systemic inflammation, neuroinflammation, and cognitive functioning in older adults.

### MIND diet and systemic inflammation

We hypothesized that MIND diet adherence would directly correlate with systemic inflammation. However, while systemic inflammation was significantly associated with adherence to MIND-discouraged food groups specifically, no association was observed with total MIND diet adherence. Previous studies on MIND diet adherence and systemic inflammation (18–20) showed inconsistent results. Our findings align with the study of Sager et al., showing no direct association between MIND diet adherence and the systemic inflammation markers CRP and IL-6 (20). Compared to our population, this study showed similar median MIND (8.5) and CRP (1.50) scores. Our findings are not consistent with the studies of Fei et al. and Liu et al., who found associations between MIND diet adherence and various systemic inflammation markers, including CRP (18,19), WBC (18), and neutrophils (18). In both studies, average MIND diet adherence was substantially lower compared to our sample whereas systemic inflammation markers were higher. To illustrate, Fei et al. reported a MIND diet median and IQR of 7.0 [5.5, 8.5], CRP median and IQR of 2.3 [1.1, 4.8], and mean WBC and NLR levels of 6.7 and 2.2, respectively (18). Liu et al. reported mean MIND diet scores of 6.60L±L2.27 and CRP levels of 2.88L±L1.48 (19). In other words, despite our at-risk selection criteria, the overall health status of our study population may have been too high to detect robust associations, which could explain the currently observed peripheral-central effects at lower MIND scores (≤7.0) only.

Moreover, Fei and colleagues showed that the relation between MIND diet adherence and systemic inflammation (specifically CRP, WBC and neutrophils) was stronger in men and in individuals with lower education (17). Both of these groups are relatively underrepresented in our study population.

Notably, we observed a specific association between adherence to MIND-discouraged food groups and systemic inflammation. This finding suggests that limiting discouraged food groups (such as saturated fats, sugars, and processed meats) might exert a greater influence on systemic inflammation than increasing consumption of recommended food groups. However, given that decomposing the overall MIND adherence score into recommended and discouraged food groups is not yet a validated approach, we encourage future studies to examine this relationship in greater detail.

### MIND diet and neuroinflammation

Based on the limited available evidence (38,39), we hypothesized a negative correlation between MIND diet adherence and neuroinflammation. However, this association was not observed in our study. Remarkably, the only prior study assessing MIND diet adherence and (post-mortem) neuroinflammation (38) demonstrated effects specifically in the hippocampus, a brain region known for its vulnerability to neuroinflammation in ageing (27). Despite acquiring hippocampal ¹H-MRS spectra, examination of neuroinflammation in this region was not feasible due to inadequate spectral quality. This poor spectral quality was likely attributable to suboptimal signal-to-noise ratio inherent to the 3 tesla field strength for deep brain spectroscopy in combination with our older population (50). We therefore recommend that future studies assess ¹H-MRS-based neuroinflammation across multiple brain regions (including the hippocampus) rather than solely in the dlPFC, utilizing higher field strength MR scanners (e.g., 7 tesla systems) offering more optimal signal-to-noise ratios (51). Furthermore, while ¹H-MRS represents a valuable approach for measuring brain metabolites in vivo and non-invasively, it remains an indirect proxy for neuroinflammation (52). Additionally, although we selected the three metabolites most consistently associated with neuroinflammation in prior research (24,53), consensus regarding the optimal metabolic markers of neuroinflammation remains lacking. Future studies could complement or validate these ¹H-MRS findings with more direct neuroinflammatory measures, such as translocator protein (TSPO) positron emission tomography (PET) imaging or cerebrospinal fluid (CSF) inflammatory biomarkers (54,55). Overall, our results emphasize that the relation between MIND diet adherence and neuroinflammation may be more complex, topographical, and subtle than initially hypothesized, especially given our cognitively healthy population.

Importantly, we were the first to examine the role of MIND diet adherence in the association between systemic inflammation and neuroinflammation, demonstrating that MIND diet adherence moderated this relationship. Specifically, in individuals with lower adherence to the MIND diet (identified as scores ≤7), systemic inflammation was positively associated with neuroinflammation. Therefore, we propose that MIND dietary components likely modulate the peripheral-to-central inflammatory pathway, although for this cross-sectional analysis we cannot conclude any directionality. A crucial gatekeeper within this peripheral-to-central pathway is the BBB. Given that dietary factors may both protect and compromise BBB integrity and function (56), we suggest that the BBB might be a key player in MIND diet’s moderation of the link between systemic- and neuroinflammation. Supporting this mechanism, animal studies consistently demonstrate that Western-style and high-fat diets compromise BBB integrity and increase permeability in rodent models (57–59), though human studies are lacking. Moreover, a human RCT demonstrated that a 6 months of extra-virgin olive oil intervention – a key MIND diet component – reduced BBB permeability in individuals with mild cognitive impairment (60). Future studies should investigate this potential role of the BBB as a critical mediator in the link between MIND dietary components and the peripheral-to-central inflammatory cascade. Advanced in vivo neuroimaging techniques to assess BBB permeability in humans, such as gadolinium-enhanced MRI and diffusion-weighted arterial spin labeling, could provide crucial insights (61).

### MIND diet and cognition

In contrast to our expectations, we found no association between MIND diet adherence and cognitive functioning. Although the majority of previous observational studies have demonstrated associations between MIND diet adherence and cognition (4), our study did not replicate these findings. As previously discussed, the relatively healthy and highly educated profile of our population may have restricted detectable variation, potentially extending to cognitive outcomes as well. Nevertheless, direct comparison of cognitive statuses across studies was hampered by substantial heterogeneity in cognitive assessment methods. Additionally, most studies included in the meta-analysis by van Soest et al. (4) used the original US-based MIND diet scoring, whereas we used a version adapted to the Dutch context and serving sizes (MIND-NL) (8), which may partially account for our divergent findings.

Notably, we again observed a moderating effect of MIND diet adherence on the translation of systemic inflammation to lower cognitive functioning (with MIND scores ≤7 as cut-off). This moderating effect suggests that the relation between MIND diet and cognition may be more nuanced than currently understood and could potentially explain the inconsistent results of prior experimental trials and longitudinal studies compared with observational cross-sectional studies. Moreover, as this moderating pattern is similar to the observed moderation of the relation between systemic- and neuroinflammation by MIND diet adherence, we suggest that these pathways might be linked. However, we could not identify a mediating effect of neuroinflammation, as neuroinflammation and cognition were not significantly correlated. Importantly, mediation analyses on cross-sectional data should be interpreted with caution given the lack of causal inference (62). Future confirmatory, longitudinal studies with larger populations should therefore examine this potential mediation. Moreover, it should be noted that cognitive decline is a complex process that relies on multiple underlying mechanisms (such as cerebral perfusion and protein accumulation), with neuroinflammation constituting one important element in this multifaceted process (63,64). Finally, we hypothesize that the BBB might also play a crucial role in this peripheral-central moderating effect of MIND diet components on cognition (65,66).

### Gut-immune-brain axis: role of intestinal barrier permeability

We hypothesized that IB permeability would partially mediate the effects of MIND diet components on systemic inflammation. Although MIND diet adherence was not directly associated with IB permeability, we observed a moderate correlation between IB permeability and systemic inflammation, driven primarily by zonulin and LBP. This finding aligns with prior research in (older) adults which demonstrated a relation between IB permeability and systemic inflammation (67,68), and identified zonulin and LBP as the permeability markers most strongly linked to metabolic health and inflammatory status (67). Consistent with the patterns we observed for systemic inflammation, neuroinflammation, and cognition, the IB permeability-neuroinflammation relation was significantly moderated by MIND diet adherence. This association was only evident in individuals with lower adherence (scores ≤7.5). Moreover, we demonstrated that systemic inflammation mediated the IB permeability-neuroinflammation relation exclusively in the individuals with lower MIND diet adherence (based on MIND score median split ≤8.75). In other words, higher MIND diet adherence might protect against the systemic inflammation-mediated relationship between IB permeability and neuroinflammation, although longitudinal studies should confirm such a protective effect. Although the mediation effect itself was robust, the relatively small subsample (based on median split) precluded definitive determination of full versus partial mediation, warranting replication in larger cohorts.

Importantly, the IB and BBB show structural similarities and their integrity appears linked, as biomarkers of IB and BBB permeability correlate in various conditions (69–71). Given that we propose the BBB as a potential target of the MIND diet, observed correlations with IB permeability markers may indicate concurrent BBB involvement, although this warrants further investigation.

## Conclusions

Collectively, our findings suggest that higher adherence to the MIND diet – a Mediterranean-type diet with emphasis on green leafy vegetables and berries – may protect against the detrimental link between systemic inflammation and both neuroinflammation and cognitive dysfunction in aging. Moreover, we demonstrated that greater adherence to the MIND diet may specifically be protective against the systemic inflammation-mediated relationship between intestinal barrier permeability and neuroinflammation. These results implicate that MIND diet components can interfere with inflammatory pathways implicated in cognitive functioning and advance our mechanistic understanding of the gut-immune-brain axis in ageing. A MIND diet adherence score of at least 7.5 to 8.0 was robustly identified as a threshold for protective effects, providing clinicians with a concrete target for evidence-based dietary counseling, even though higher scores remain preferable for optimal benefits. Future intervention studies should confirm these associations and investigate the underlying mechanisms, for which we suggest additional focus to be put on blood-brain barrier integrity measures.

## Data Availability

Data available upon request.
Data will become available online at Radboud Data Repository after publication.

## List of abbreviations

1H-MRS: Proton magnetic resonance spectroscopy
AD: Alzheimer’s Disease
BBB: Blood-brain barrier
BMI: Body mass index
CHESS: Chemically selective water suppression
CRP: C-reactive protein
CSF: Cerebrospinal fluid
CVD: Cardiovascular disease
DASH: Mediterranean-Dietary Approaches to Stop Hypertension
dlPFC: Dorsolateral prefrontal cortex
DST: Digit Span Test
DSST: Digit Symbol Substitution Test
FFQ: Food Frequency Questionnaire
FWHM: Full width at half maximum
IB: Intestinal barrier
IL: Interleukin
IQR: Interquartile range
LBP: Lipopolysaccharide Binding Protein
MIND: Mediterranean-Dietary Approaches to Stop Hypertension Intervention for Neurodegenerative Delay
MoCA: Montreal Cognitive Assessment
MP2RAGE: Magnetization-prepared 2 rapid gradient echo
MRI: Magnetic resonance imaging
NLR: Neutrophil-to-lymphocyte ratio
NTB: Neuropsychological test battery
PCA: Principal component analysis
PC1: First principal component
PET: Positron Emission Tomography
PRESS: Point RESolved Spectroscopy
RAVLT: Rey Auditory Verbal Learning Test
SNR: Signal-to-noise ratio
SPM12: Statistical Parametric Mapping, version 12
TE: Echo time
TICS-M1: Telefonisch Interview voor Cognitieve Status (TICS-M) version 1
TMT: Trail Making Test
TR: Repetition time
VFT: Verbal Fluency Test
WBC: White blood cell

## Declarations

### Ethics approval and consent to participate

The HELI study was conducted in compliance with the Declaration of Helsinki for research involving human participants and the Medical Research Involving Human Subjects Act (WMO; ‘Wet Medisch-wetenschappelijk Onderzoek met mensen’). The complete procedure was approved by the local Ethics Committee (METC Oost-Nederland, NL78263.091.21) on December 2^nd^ 2022 and registered in the Clinical Trial Register (ClinicalTrials.gov ID: NCT05777863). Written informed consent was obtained from each participant during the first study visit.

### Consent for publication

Not applicable.

### Availability of data and materials

Data supporting the findings of this study are available from the Radboud Data Repository via the following URL: [URL].

### Authors’ contributions

LR, JO and EA conceptualized this study. MvT and JM were involved in design, analyses and interpretation of the blood-based biomarkers. LR, IdL and NP were involved in the ^1^H-MRS data analysis and/or data quality assessment. LR conducted the statistical analysis. JO and EA guided the statistical analysis and contributed to the interpretation of results. MvL, LR, MvT, IdL, JM, JO and EA were involved in the design and data collection of the HELI study. All authors read and approved the final manuscript.

## Acknowledgements

To start with, we would like to express our gratitude to all the participants of the HELI study for volunteering. Moreover, we would like to thank Diede Booltink for her practical contributions in MRI scanning; Paul Gaalman for his assistance in the neuroimaging lab and his help in setting up the MRI protocol; Viola Hollestein and Jilly Naaijen for their advice on ^1^H-MRS analyses; and Els Oosterink and Monic Tomassen for conducting the permeability marker blood analyses. Furthermore, we would like to express our gratitude to all MSc thesis students that assisted within the HELI study for their practical contributions, in particular Geert Woort who conducted the first steps within this analysis. Finally, we extend our appreciation to the Human Nutrition Research Unit team and the DCCN research assistant pool for their support in the practical execution of the study.

