## Supplement for "MIND-NL diet adherence moderates the relation of low-grade systemic inflammation with neuroinflammation and cognitive functioning: an exploratory cross-sectional study in older adults"

### Supplementary tables

**Supplementary Table 1.** Modifiable cardiovascular risk factor scale used for study inclusion

| Risk Factor (self-reported) | Point(s) |
|---|---|
| BMI ≥ 25kg/m$^2$ (overweight) | 1 |
| Physical inactivity (below the 2020 WHO guidelines: <300 minutes of moderate intensity aerobic physical activity or <150 minutes of vigorous intensity aerobic physical activity per week, spread out over several days) | 1 |
| Hypertension (systolic blood pressure ≥140 mmHg, and diastolic blood pressure ≥90 mmHg) | 1<br>(2 points are assigned if hypertension is not being actively treated with antihypertensive medication, given the increased cardiovascular burden) |
| Hypercholesterolemia (total cholesterol >5 mmol/L, or LDL-cholesterol >3 mmol/L) | 1 |
| Diabetes type-II | 1 |
| Mild cardiovascular disease (e.g. intermittent claudication, varicose veins; In contrast, moderate or severe cardiovascular disease such as stroke, angina pectoris, heart failure, myocardial infarction or revascularization surgery in the last 12 months before pre-screening are exclusion criteria) | 1 |

**Supplementary Table 2.** Components of the Dutch version of the Mediterranean-DASH Diet Intervention for Neurodegenerative Delay (MIND-NL) diet and cut-off values per score type.

| **MIND-NL food groups** | **Score** | | |
|---|---|---|---|
| | **0** | **0.5** | **1** |
| Green leafy vegetables | ≤29 g/day | >29 - <100 g/day | ≥100 g/day |
| Other vegetables | <71 g/day | ≥ 71 -<100 g/day | ≥100 g/day |
| Berries and strawberries | <7 g/day | ≥7 - <36 g/day | ≥36 g/day |
| Legumes | <9 g/day | ≥9 - <26 g/day | ≥26 g/day |
| Nuts | <3 g/day | ≥3 - <20 g/day | ≥20 g/day |
| Fish (not fried) | <13 g/day | 13 g/day | >13 g/day |
| Whole grains | <30 g/day | ≥30 - <90 g/day | ≥90 g/day |
| Poultry | <14 g/day | ≥14 - <29 g/day | ≥29 g/day |
| Olive oil | <15 g/day | ≥15 - <30 g/day | ≥30 g/day |
| Butter and stick margarine | ≥10 g/day | >5 - <10 g/day | ≤5 g/day |
| Full-fat cheese | ≥30 g/day | >9 - <30 g/day | ≤9 g/day |
| Red and processed meat | ≥100 g/day | >43 - <100 g/day | ≤43 g/day |
| Wine | >100 ml/d | NA | ≤100 ml/d |
| Take out, fried foods and snacks | >3 serving eq/wk | >1 - ≤3 serving eq/wk | ≤1 serving eq/wk |
| Cookies, pastries and sweets | >4 serving eq/wk | >2 - ≤4 serving eq/wk | ≤2 serving eq/wk |

**Supplementary Table 3.** Data quality measures and tissue fractions of [1]H-MRS data (*n*=88) within the dorsolateral prefrontal cortex.

| | Cr SNR | Cr FWHM | Water FWHM | relResA | GM % | WM % | CSF % |
|---|---|---|---|---|---|---|---|
| ***Mean ± SD*** | 62.97 ± 11.0 | 7.87 ± 1.72 | 8.52 ± 1.85 | 2.92 ± 1.8 | 34 ± 6 | 61 ± 8 | 4.9 ± 3.7 |

*Cr = Creatin; SNR = signal-to-noise ratio; FWHM = full width at half maximum; GM = grey matter; WM = white matter; CSF = cerebrospinal fluid; SD = standard deviation*

**Supplementary Table 4.** Linear model statistics with and without interaction per model

| Models with terms | Estimate | CI (lower) | CI (upper) | Std. Error | t value | Pr(>\|t\|) |
|---|---|---|---|---|---|---|
| **MODEL 1a: Neuroinflammation ~ age + sex + relResA + MIND adherence + systemic inflammation (PC1)** | | | | | | |
| ***n*=88** | | | | | | |
| *$R^2$ = 0.023; Adj. $R^2$ = -0.037; F-statistic = 0.39; P = 0.86* | | | | | | |
| (Intercept) | 1.30 | -3.40 | 5.99 | 2.36 | 0.55 | 0.58 |
| Age | -0.02 | -0.09 | 0.05 | 0.04 | -0.57 | 0.57 |
| Sex (male) | -0.09 | -0.77 | 0.59 | 0.34 | -0.26 | 0.80 |
| relResA (spectral quality) | 0.06 | -0.12 | 0.23 | 0.09 | 0.65 | 0.52 |
| MIND adherence | 0.00 | -0.16 | 0.16 | 0.08 | -0.04 | 0.97 |
| Systemic inflammation (PC1) | 0.12 | -0.11 | 0.36 | 0.12 | 1.05 | 0.30 |
| **MODEL 1b: Neuroinflammation ~ age + sex + relResA + MIND adherence + systemic inflammation (PC1) + MIND adherence:systemic inflammation (PC1)** | | | | | | |
| ***n*=88** | | | | | | |
| *$R^2$ = 0.074; Adj. $R^2$ = 0.006; F-statistic = 1.09; P = 0.38* | | | | | | |
| (Intercept) | 1.39 | -3.21 | 5.99 | 2.31 | 0.60 | 0.55 |
| Age | -0.03 | -0.10 | 0.05 | 0.04 | -0.73 | 0.47 |
| Sex (male) | -0.10 | -0.76 | 0.57 | 0.33 | -0.29 | 0.77 |
| relResA (spectral quality) | 0.07 | -0.10 | 0.24 | 0.09 | 0.84 | 0.41 |
| MIND adherence | 0.02 | -0.14 | 0.17 | 0.08 | 0.23 | 0.82 |
| Systemic inflammation (PC1) | 1.11 | 0.16 | 2.05 | 0.48 | 2.32 | 0.02 |
| MIND adherence: Systemic inflammation (PC1) | -0.11 | -0.22 | -0.01 | 0.05 | -2.12 | 0.04 |
| **MODEL 2a: Cognition ~ age + sex + education + MIND adherence + systemic inflammation (PC1)** | | | | | | |
| ***n*=88** | | | | | | |
| *$R^2$ = 0.21; Adj. $R^2$ = 0.16; F-statistic = 4.36; P = 0.001* | | | | | | |
| (Intercept) | 3.41 | 1.70 | 5.13 | 0.86 | 3.95 | 0.00 |
| Age | -0.05 | -0.08 | -0.03 | 0.01 | -4.25 | 0.00 |
| Sex (male) | -0.02 | -0.24 | 0.21 | 0.11 | -0.13 | 0.89 |
| Education level | 0.02 | -0.05 | 0.08 | 0.03 | 0.47 | 0.64 |
| MIND adherence | 0.01 | -0.04 | 0.06 | 0.03 | 0.32 | 0.75 |
| Systemic inflammation (PC1) | -0.02 | -0.10 | 0.05 | 0.04 | -0.61 | 0.55 |
| **MODEL 2b: Cognition ~ age + sex + education + MIND adherence + systemic inflammation (PC1) + MIND adherence:systemic inflammation (PC1)** | | | | | | |
| ***n*=88** | | | | | | |
| *$R^2$ = 0.27; Adj. $R^2$ = 0.21; F-statistic = 4.89; P <0.001* | | | | | | |
| (Intercept) | 3.33 | 1.66 | 4.99 | 0.84 | 3.97 | 0.00 |
| Age | -0.05 | -0.08 | -0.03 | 0.01 | -4.19 | 0.00 |
| Sex (male) | -0.02 | -0.24 | 0.20 | 0.11 | -0.16 | 0.87 |
| Education level | 0.02 | -0.04 | 0.08 | 0.03 | 0.63 | 0.53 |
| MIND adherence | 0.00 | -0.05 | 0.05 | 0.03 | 0.03 | 0.98 |
| Systemic inflammation (PC1) | -0.40 | -0.71 | -0.09 | 0.16 | -2.56 | 0.01 |
| MIND adherence: Systemic inflammation (PC1) | 0.04 | 0.01 | 0.08 | 0.02 | 2.48 | 0.02 |

| | | | | | | |
|---|---|---|---|---|---|---|
| **MODEL 3a: Neuroinflammation ~ age + sex + relResA + MIND adherence + intestinal barrier permeability (PC1)** | | | | | | |
| ***n*=71** | | | | | | |
| *$R^2$ = 0.028; Adj. $R^2$ = -0.047; F-statistic = 0.37; P = 0.87* | | | | | | |
| (Intercept) | 1.04 | -4.45 | 6.52 | 2.75 | 0.38 | 0.71 |
| Age | -0.02 | -0.10 | 0.07 | 0.04 | -0.42 | 0.68 |
| Sex (male) | -0.13 | -0.91 | 0.65 | 0.39 | -0.32 | 0.75 |
| relResA (spectral quality) | 0.08 | -0.10 | 0.27 | 0.09 | 0.89 | 0.38 |
| MIND adherence | -0.01 | -0.19 | 0.17 | 0.09 | -0.11 | 0.91 |
| Intestinal barrier permeability (PC1) | 0.15 | -0.16 | 0.46 | 0.15 | 0.96 | 0.34 |
| **MODEL 3b: Neuroinflammation ~ age + sex + relResA + MIND adherence + intestinal barrier permeability (PC1) + MIND adherence: intestinal barrier permeability (PC1)** | | | | | | |
| ***n*=71** | | | | | | |
| *$R^2$ = 0.083; Adj. $R^2$ = -0.003; F-statistic = 0.97; P = 0.46* | | | | | | |
| (Intercept) | 1.09 | -4.28 | 6.46 | 2.69 | 0.41 | 0.69 |
| Age | -0.02 | -0.11 | 0.06 | 0.04 | -0.58 | 0.56 |
| Sex (male) | 0.18 | -0.65 | 1.00 | 0.41 | 0.43 | 0.67 |
| relResA (spectral quality) | 0.09 | -0.09 | 0.28 | 0.09 | 1.00 | 0.32 |
| MIND adherence | 0.02 | -0.16 | 0.20 | 0.09 | 0.21 | 0.83 |
| Intestinal barrier permeability (PC1) | 1.71 | 0.09 | 3.32 | 0.81 | 2.11 | 0.04 |
| MIND adherence: Intestinal barrier permeability (PC1) | -0.17 | -0.34 | 0.00 | 0.09 | -1.97 | 0.05 |

**Supplementary Table 5. Mediation model statistics predicting cognition**. SEM-based mediation analysis predicting mediating effect of neuroinflammation within relation between MIND diet adherence:systemic inflammation interaction and cognition (*n*=88).

**Mediation model statistics**

**Path Coefficients**

| *Path* | *Coefficient (SE)* | *95% bootstrap CI* | *P-value* |
|---|---|---|---|
| **a**: MIND adherence × Systemic inflammation (PC1) → Neuroinflammation (PC1) | -0.115 ( 0.068 ) | [-0.269, -0.004] | 0.09 |
| **b**: Neuroinflammation (PC1) → Cognition composite score (Z) | -0.028 ( 0.043 ) | [-0.127, 0.041] | 0.51 |
| **c'**: MIND adherence × Systemic inflammation (PC1) → Cognition composite score (Z) (direct) | 0.040 ( 0.023 ) | [-0.011, 0.081] | 0.09 |

**Effect Estimates**

| *Effect type* | *Estimate (SE)* | *95% bootstrap CI* | *P-value* |
|---|---|---|---|
| Direct effect (c’) | 0.040 ( 0.023 ) | [-0.011, 0.081] | 0.09 |
| Indirect effect (a x b) | 0.003 ( 0.006 ) | [-0.005, 0.019] | 0.58 |
| Total effect (c) | 0.043 ( 0.022 ) | [-0.002, 0.084] | 0.05 |

**Supplementary Table 6. Mediation model statistics predicting neuroinflammation**. SEM-based mediation analysis predicting mediating effect of systemic inflammation within relation between intestinal barrier permeability and neuroinflammation within the low (*n*=35) and high (*n*=36) MIND diet adherence group based on median split.

| **Mediation model statistics** | **Low MIND group (≤8.75, *n*=35)** | | | **High MIND group (>8.75, *n*=36)** | | |
|---|---|---|---|---|---|---|
| **Path Coefficients** | | | | | | |
| ***Path*** | ***Coefficient (SE)*** | ***95% bootstrap CI*** | ***P-value*** | ***Coefficient (SE)*** | ***95% bootstrap CI*** | ***P-value*** |
| **a:** Intestinal permeability (PC1) → Systemic inflammation (PC1) | 0.771 ( 0.253 ) | [0.264, 1.272] | 0.002 | 0.498 ( 0.213 ) | [0.123, 0.975] | 0.02 |
| **b**: Systemic inflammation (PC1) → Neuroinflammation (PC1) | 0.553 ( 0.230 ) | [0.155, 1.055] | 0.02 | -0.129 ( 0.174 ) | [-0.518, 0.176] | 0.46 |
| **c'**: Intestinal permeability (PC1) → Neuroinflammation (PC1) (direct) | 0.329 ( 0.417 ) | [-0.391, 1.204] | 0.43 | -0.046 ( 0.142 ) | [-0.295, 0.277] | 0.75 |
| **Effect Estimates** | | | | | | |
| ***Effect type*** | ***Estimate (SE)*** | ***95% bootstrap CI*** | ***P-value*** | ***Estimate (SE)*** | ***95% bootstrap CI*** | ***P-value*** |
| Direct effect (c') | 0.329 ( 0.417 ) | [-0.391, 1.204] | 0.43 | -0.046 ( 0.142 ) | [-0.295, 0.277] | 0.75 |
| Indirect effect (a x b) | 0.427 ( 0.212 ) | [0.072, 0.891] | 0.04 | -0.064 ( 0.096 ) | [-0.298, 0.081] | 0.51 |
| Total effect (c) | 0.756 ( 0.493 ) | [-0.125, 1.716] | 0.13 | -0.110 ( 0.134 ) | [-0.379, 0.160] | 0.41 |

### Supplementary figures

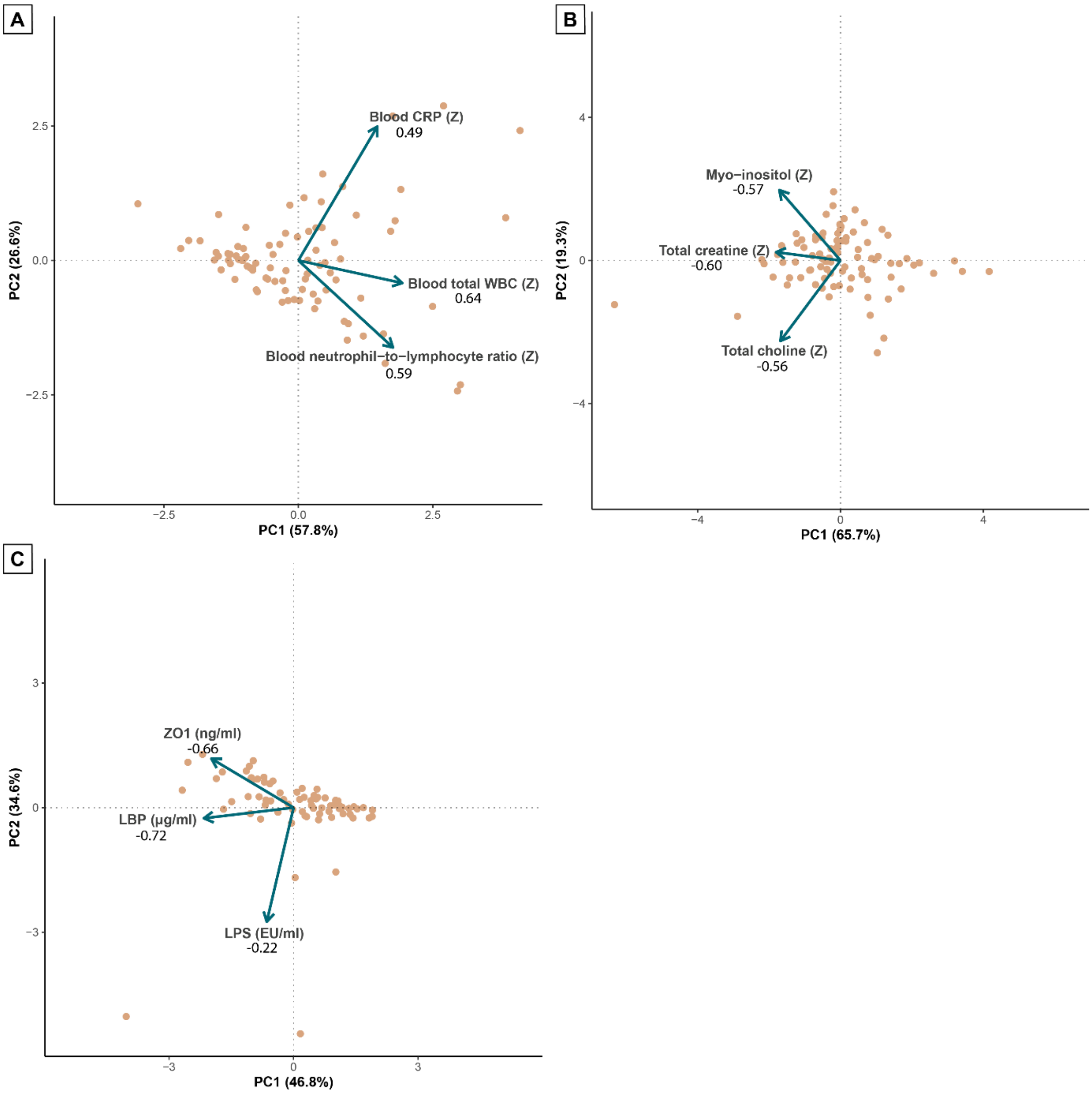


***Supplementary Figure 1. PCA plots***. *Principal Component Analysis plots of systemic inflammation (n=88) (**A**), neuroinflammation (n=88) (**B**), and intestinal barrier permeability (n=71) (**C**). PC1 loadings are shown below each variable. LPS PC1 loadings were lower on compared to zonulin and LBP for intestinal barrier permeability, probably due to a few extreme outliers.*

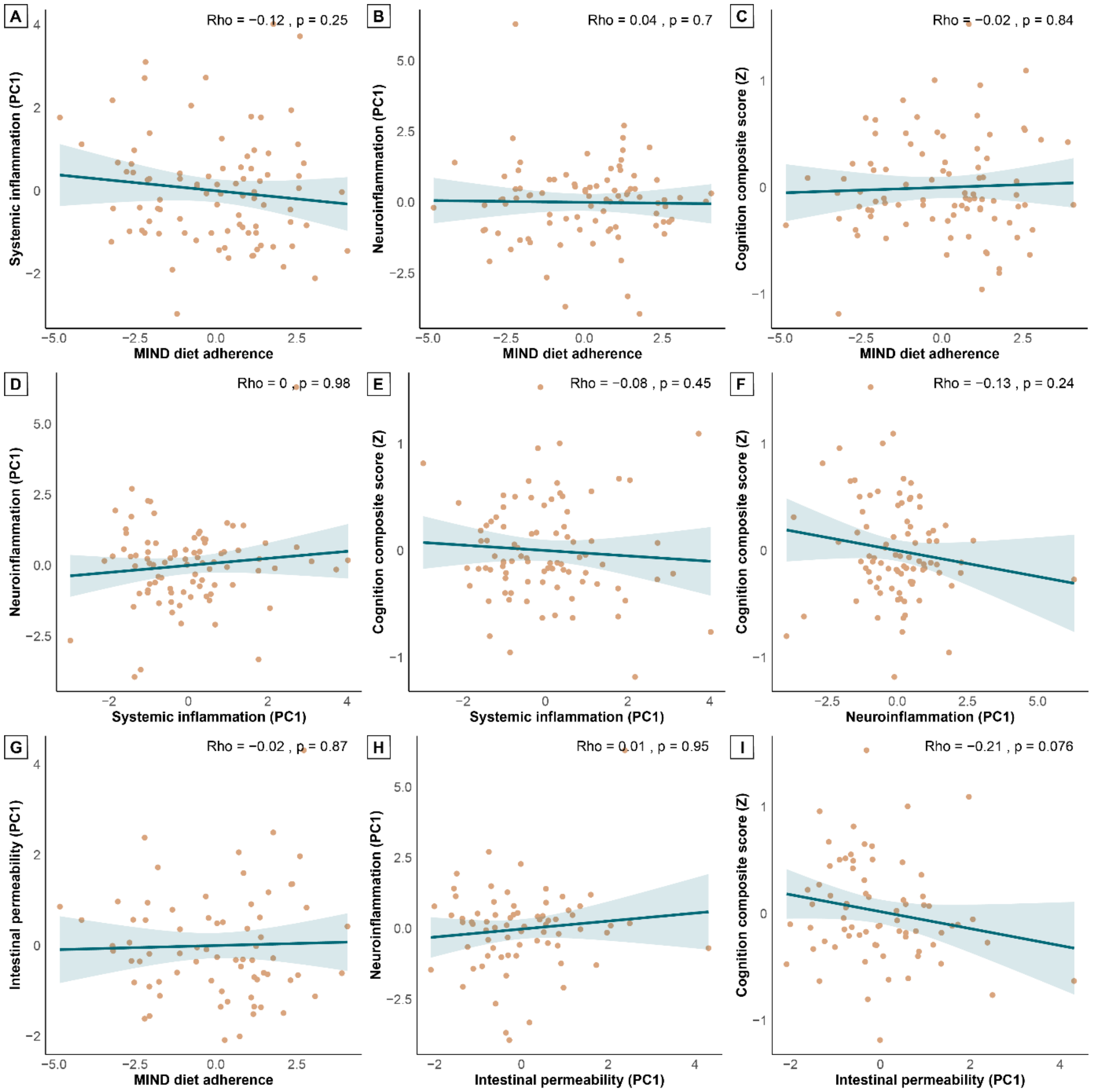


***Supplementary Figure 2. Pairwise Spearman correlation scatterplots for MIND diet adherence, systemic inflammation, neuroinflammation, cognition, and intestinal barrier permeability.*** *Scatterplots showing correlations between MIND diet adherence and systemic inflammation (n=88) (**A**), MIND diet adherence and neuroinflammation (n=88) (**B**), MIND diet adherence and cognition composite score (n=88) (**C**), systemic inflammation and neuroinflammation (n=88) (**D**), systemic inflammation and cognition composite score (n=88) (**E**), neuroinflammation and cognition composite score (n=88) (**F**), MIND diet adherence and intestinal barrier permeability (n=71) (**G**), intestinal barrier permeability and neuroinflammation (n=71) (**H**), and intestinal barrier permeability and cognition composite score (n=71) (**I**).*

*Orange dots represent individual subjects. Trendline and confidence interval are displayed in blue. Spearman's rho and p-value are reported top right. Data was corrected for age, sex, relResA/spectral quality (neuroinflammation), and education levels (cognition). Residuals were used for correlation analyses*.

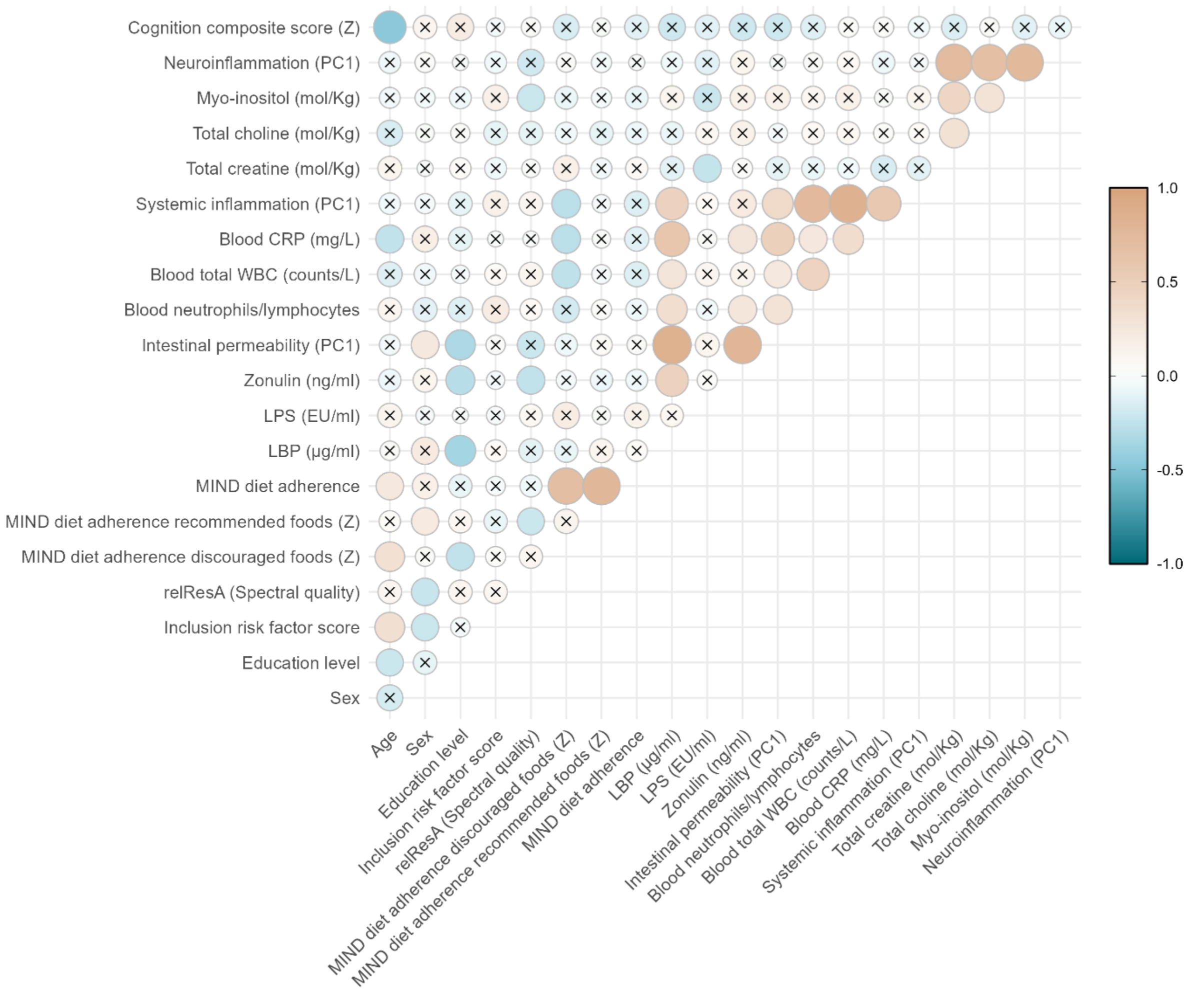


***Supplementary Figure 3. Spearman correlation heatmap of all variables, subscores and covariates (n=88).*** *Heatmap showing correlations between MIND diet adherence, intestinal barrier permeability, systemic inflammation, neuroinflammation, cognition, subscores for each combination score, and other variables of interest/covariates. Orange colours indicate a positive correlation; blue colours indicate a negative correlation. Circle size and color intensity indicate strength of the correlation (Spearman's rho). Absence of a cross indicate significance (p≤0.05) after FDR correction. For intestinal barrier permeability and subscores, correlations include n=71 participants. FDR = false discovery rate.*

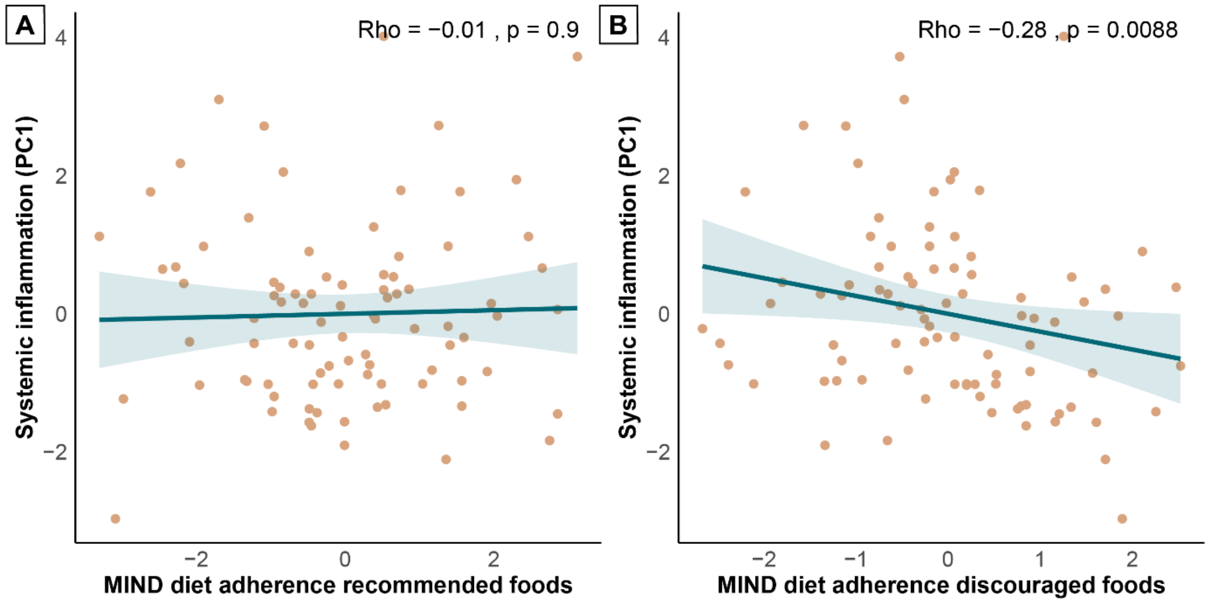


***Supplementary Figure 4. Spearman correlation scatterplots for MIND diet adherence to recommended and discouraged foods, and systemic inflammation (n=88).*** *Scatterplots showing correlations between MIND diet adherence to (consuming) recommended foods and systemic inflammation (**A**), and MIND diet adherence to (limiting) discouraged foods and systemic inflammation (**B**). Orange dots represent individual subjects. Trendline and confidence interval are displayed in blue. Spearman's rho and p-value are reported top right. Data was corrected for age and sex. Residuals were used for correlation analyses.*

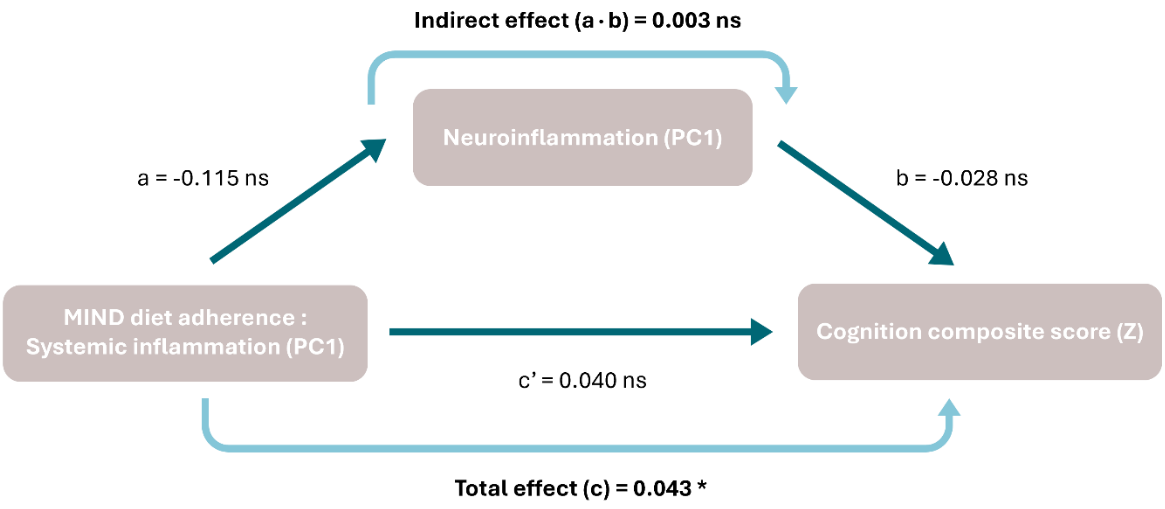


***Supplementary Figure 5. Mediation analysis diagram for MIND adherence:systemic inflammation interaction, neuroinflammation, and cognition (n=88).** Diagram showing the mediating role of neuroinflammation in the relation between the interaction term MIND adherence:systemic inflammation and cognition. Significance is indicated with asterisks (*** = 0.001, ** = 0.01, * = 0.05, ns = non-significant).*
